# Trajectories of Depressive Symptoms Across the Menopause Transition: A Longitudinal Analysis from a Prospective UK Cohort Study

**DOI:** 10.64898/2026.04.27.26351577

**Authors:** Rochelle Knight, Carol Joinson, Abigail Fraser, Ana Gonçalves Soares

## Abstract

**Background:** Women are twice as likely to experience depression compared with men. Midlife is a period of increased vulnerability to depression in women and is a stage that coincides with the menopausal transition. Although prospective studies suggest that depressive symptoms increase during this period, findings are inconsistent and it remains difficult to disentangle the effects of reproductive aging from chronological aging. We examined trajectories of depressive symptoms across midlife in relation to reproductive and chronological aging and assessed the association between menopause stages and depression.

**Methods:** We analysed data from 2,036 women from a UK community-based pregnancy cohort followed over three decades from the early 1990’s to early 2020’s. Depressive symptoms were assessed at up to 11 timepoints using the Edinburgh Postnatal Depression Scale (EPDS), with depression defined as a score ≥ 13. Linear mixed effects models were used to characterise trajectories of depressive symptoms across reproductive and chronological age, and to estimate associations between menopause stages and depression, adjusting for ethnicity, social class, age at menarche, educational attainment, material hardship, social support, smoking status, body mass index (BMI), and alcohol intake.

**Results:** Depressive symptoms increased gradually with both reproductive and chronological age; however, there was little evidence of a marked increase in levels of depressive symptoms during the peri- or postmenopause stages. Compared with the reproductive period, perimenopause was associated with higher odds of depression (OR 1.19; 95% CI 1.03 to 1.39), whereas there was no evidence of increased odds during postmenopause (OR 0.99; 95% CI 0.83 to 1.19).

**Conclusions:** Overall, reproductive ageing appears to contribute relatively little to long-term changes in depressive symptoms beyond the broader effects of chronological ageing. The odds of depression were highest in the perimenopause and did not persist into postmenopause, indicating that this stage may represent a window of vulnerability for a subgroup of women. Identifying such subgroups could help inform targeted prevention and support strategies.

## Background

Depression is a major global health burden, affecting approximately 15% of adults in high-income countries.(1) Women experience major depression at roughly twice the rate of men(1–3), with the greatest sex disparity observed during women’s reproductive years(4) and attenuating in later life(5). Midlife has emerged as a period of increased vulnerability to depressive symptoms in women, often coinciding with the menopausal transition.(6)

Some prospective studies have found evidence that the menopausal transition is associated with an increase in levels of depressive symptoms(7–9), although findings are inconsistent(8, 9). A meta-analysis of seven longitudinal studies (n=9,141) reported elevated odds of depression in perimenopause relative to premenopause, with no increase observed in postmenopause.(10) It has also been hypothesised that later menopause, reflecting a longer reproductive period and greater lifetime exposure to endogenous oestrogen, may reduce depression risk(5). A meta-analysis(11) of 14 studies including over 67,000 women found that both a later age at menopause and a longer reproductive period were associated with lower odds of postmenopausal depression. More recent longitudinal evidence reported that a longer reproductive period (from menarche to menopausal transition onset) was associated with reduced risk of depression during the menopausal transition and up to 10 years postmenopause.(12)

Disentangling the effects of reproductive aging – calculated as years before and after the final menstrual period (FMP) – from chronological aging is challenging, as both progress simultaneously and are highly correlated. The aims of this study are (i) to examine trajectories of depressive symptoms across midlife in relation to reproductive and chronological aging, spanning the reproductive period into postmenopause, and (ii) to assess the association between menopause stages and depression using data from a UK community-based pregnancy cohort.

## Methods

### Participants

This study used data from the mothers cohort of the Avon Longitudinal Study of Parents and Children (ALSPAC), a birth cohort that recruited pregnant women residing in Avon, UK, with an expected date of delivery between April 1991 and December 1992. The initial ALSPAC sample included 14,541 pregnancies from 14,203 unique mothers, resulting in 14,062 live births. An additional 630 mothers joined through later recruitment, bringing the total to 14,833 unique mothers (known as Generation 0 or G0). Since enrolment, participants and their families have been followed through questionnaires, research clinic assessments, and data linkage. Since 2014 study data have been collected and managed using REDCap (Research Electronic Data Capture), a secure, web-based platform hosted at the University of Bristol(13). REDCap is specifically designed to support data capture and management in research studies.

The cohort’s recruitment, representativeness, and profile have been extensively described elsewhere(14–17). A fully searchable data dictionary and variable search tool are available on the ALSPAC website (http://www.bristol.ac.uk/alspac/researchers/our-data).

The primary analysis included women who had at least one depressive symptom measure and an estimated age at natural menopause. Women with an estimated age at menopause younger than 40 years (likely reflecting premature ovarian insufficiency)—or older than 60 (considered implausible) were excluded (N= 75). As we were interested in changes in depressive symptoms associated with the natural menopause, data were censored following reports of hysterectomy, oophorectomy, or 12 months of amenorrhea attributed to surgery. This resulted in a sample of 2,036 women with 18,466 depressive symptom measures. Because it was not possible to estimate age at natural menopause for all participants, we conducted a secondary analysis that included all women with at least one depressive symptom measure and information on menopause stage, regardless of whether they had an estimated age at menopause. In this broader sample, women were classified as reproductive, perimenopausal, and postmenopausal based on the STRAW(18) criteria (N = 4,946 with 27,661 measures), as described in more detail below. The flowchart showing the derivation of the samples for both analyses is illustrated in Figure 1.

**Figure 1.**
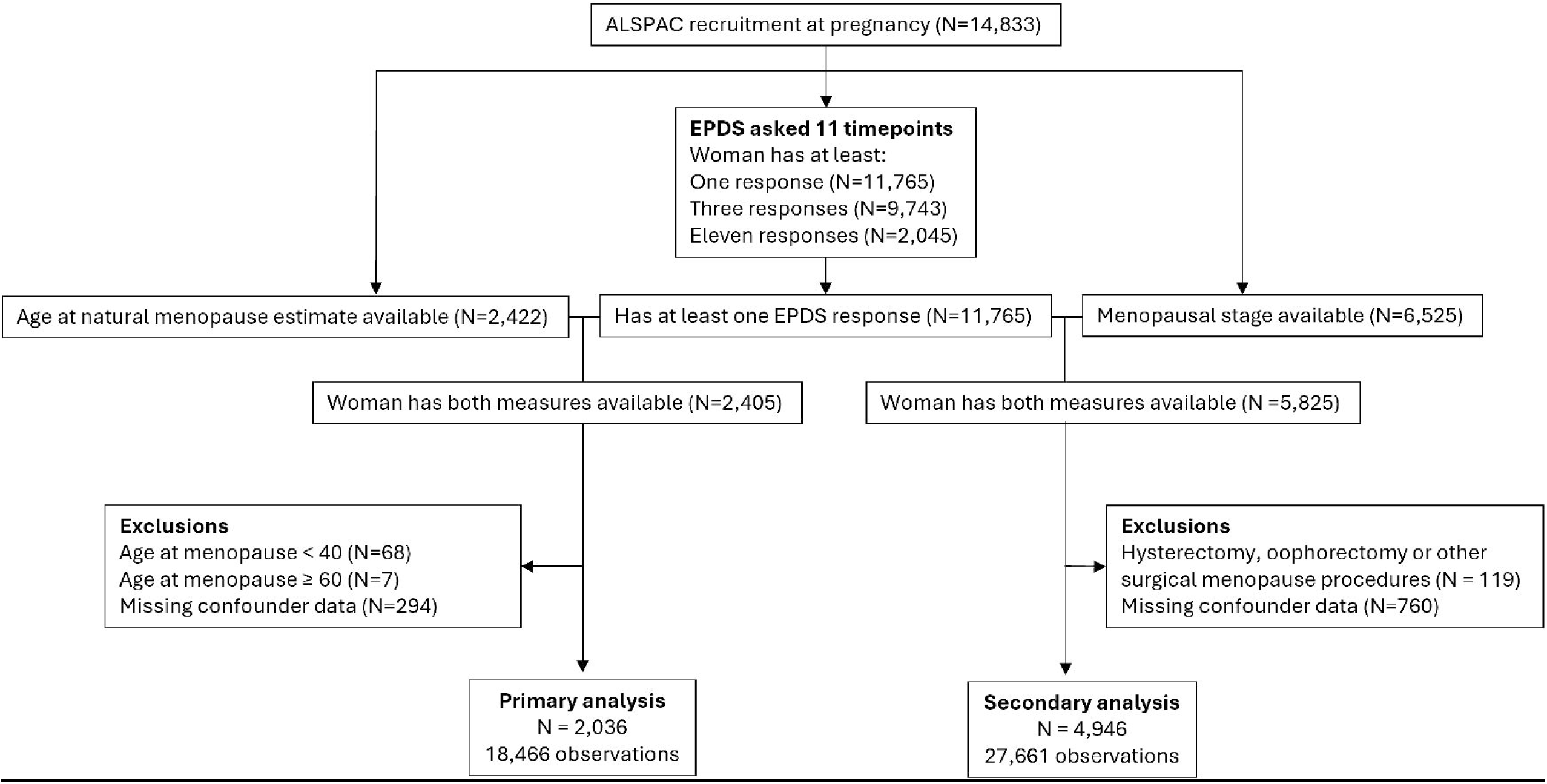
Flowchart of participants.

### Depressive symptoms

Depressive symptoms were assessed using the Edinburgh Postnatal Depression Scale (EPDS)(19). This 10-item scale assesses depressive symptoms over the past week, generating a score between 0 and 30, with higher scores indicating greater symptom severity. Although originally designed for perinatal use, the EPDS has been widely validated in general populations and is frequently employed to assess depressive symptoms in adults across the life course(19, 20).

Since enrolment, the EPDS has been administered at 11 timepoints outside the perinatal period of the index pregnancy, spanning 30 years of follow-up (Supplementary Table 1). Details on the derivation of EPDS scores are presented in Supplementary Table 2. In this study, the EPDS was used both as a continuous score and as a binary variable, with a cutoff of 13 or higher indicating probable depression (hereafter called depression) (19, 20).

### Reproductive and chronological age and menopause stage

Age at natural menopause was estimated based on self-reported menstrual bleeding patterns(21). In brief, women reported information on their menstrual cycles at up to eight timepoints from mean age 47.4 to mean age 60.7 years. The final menstrual period (FMP) was defined as the date of the last menstrual period followed by at least 12 months of amenorrhea. Age at menopause was defined as the age at the FMP. Details on how age of menopause was estimated are described elsewhere.(21)

Chronological age was self-reported at each timepoint in which EPDS was assessed. Reproductive age at each EPDS assessment was calculated by subtracting age at menopause from chronological age at the time of the EPDS measure. Reproductive age was expressed in years before and after the menopause; hence, 0 represents the age at the FMP, -1 one year before, and +1 one year after.

In addition, all women who provided menstrual cycle data – regardless of whether an age at menopause could be estimated – were classified into one of three mutually exclusive menopause stages at each EPDS assessment, based on menstrual cycle patterns and following the STRAW criteria(18):

i. Reproductive (peak and late reproductive age, STRAW category -4, -3b and -3a)
ii. Perimenopause (STRAW category -2, -1 and +1a)
iii. Postmenopause (STRAW category ≥ 1b (irrespective of the years since menopause))

Further details are provided in Supplementary Methods.

### Confounders

Selection of confounders was based on a review of the empirical literature to identify factors with potential causal effects on both age at menopause and depressive symptoms. All confounders were based on self-report and included chronological age, ethnicity, social class, age at menarche, educational attainment, material hardship, social support, smoking status, body mass index (BMI), and alcohol intake. Given some studies report a possible link between history of depression and age at menopause(22, 23), we additionally adjusted for history of depression prior to the first EPDS assessment in sensitivity analyses. More details on the questions used and categorisation of the confounders are presented in Supplementary Table 3.

Ethnicity, social class, age at menarche, educational attainment and history of depression were collected around the time of recruitment (mean age 28.6 years, SD 4.9). Missing responses for ethnicity, age at menarche, and education were replaced using data from subsequent questionnaires. Social class and history of depression were only recorded at baseline and therefore were replaced when missing.

Material hardship, social support, smoking status, BMI, and alcohol intake were treated as time-varying confounders. At each EPDS assessment, the most recent non-missing response prior to or at that time point was used.

### Statistical analyses

The primary outcome was depressive symptoms measured continuously using the EPDS, with secondary analysis treating depression as a binary outcome.

We used mixed-effects models to examine associations between depressive symptoms and both reproductive age and chronological age, accounting for repeated EPDS measures within women by including a woman-specific random intercept and a random slope for age. All women with at least one EPDS measure were included under the missing at random (MAR) assumption.

We fitted three separate model specifications:

i. reproductive age alone,
ii. chronological age alone, and
iii. reproductive age adjusted for chronological age.

For each specification we compared linear, quadratic, and restricted cubic spline (RCS) models to identify the best fit. Model fit was assessed using the Akaike Information Criterion (AIC), Bayesian Information Criterion (BIC), and visual inspection of fitted trajectories to assess plausibility and potential overfitting. All models were adjusted for the confounders specified above.

Primary analyses were restricted to women with a known age at menopause, which was required to calculate reproductive age (N = 2,036; 18,466 observations). In the reproductive age model where chronological age was included as a confounder, age was centred at 50 years (the median age at menopause) and modelled with linear and quadratic terms.

To examine whether associations between depressive symptoms and chronological age differed according to age at menopause, we tested an interaction between chronological age and age at menopause. Age at natural menopause is commonly categorised as early (40-44 years)(24), normal (45-55 years)(24) and late (over 55 years)(25). Because late menopause was only observed in 72 women, normal and late menopause were combined. Age at menopause was therefore categorised as early (40 – <45 years; *n* = 149) and normative/late (45 – <60; *n* = 1,887).

To maximise inclusion of women for whom age at menopause was unknown, we conducted secondary analyses examining the association between depressive symptoms and chronological age across menopause stages (reproductive, perimenopause, and postmenopause). These analyses included all women with at least one EPDS assessment for which menopause stage could be assigned at the time of assessment (N = 4,946; 27,661 observations). An interaction between menopause stage and chronological age was included to assess whether trajectories of depressive symptoms differed by menopause stage.

Analyses were repeated using the EPDS as a binary outcome (EPDS ≥13). Random-effects logistic regression models with a woman-specific random intercept were fitted and adjusted for the same confounders defined above. In addition, we fitted a random-effects logistic regression model with menopause stage as a categorical exposure to estimate the odds of depression in peri- and post-menopausal period compared with the reproductive period.

All analyses were performed in R (version 4.4.2) using MLwiN (version 3.13) via the *R2MLwiN* package (version 0.8.9) using the *runMLwiN* function.

### Sensitivity analyses

Several sensitivity analyses were conducted to assess the robustness of findings.

To assess the influence of extreme values, analyses were restricted to the central 90% of the distribution of reproductive age (5th–95th percentiles; N = 2,036 women contributing 16,881 observations).

EPDS scores were positively skewed, as it is common in community-based samples. Residuals from models using the original scale were assessed for normality; however, to examine the robustness of the findings, we repeated the analyses using log-transformed EPDS.

Since it was not possible to estimate age at menopause for all women, which can induce selection bias, we repeated the analysis of depressive symptoms across chronological age in the full sample of women with at least one EPDS measure (N = 9,277 women contributing 62,583 EPDS observations) to assess the potential impact of selection bias.

Given that hormone replacement therapy (HRT) is commonly used to alleviate menopausal symptoms such as hot flushes, night sweats and sleep disturbances(24) – factors that may influence depressive symptoms(26) – we examined whether associations between reproductive age and depressive symptoms differed by HRT use. Because HRT use was not reported at every timepoint EPDS was assessed, women were categorised as “ever” versus “never” users across the study period. Effect modification by HRT use was assessed by including an interaction term between HRT and reproductive age.

Finally, a history of depression was additionally included as a confounder in all main analysis models.

## Results

Among the 2,036 women included in the main analysis, the median age at menopause was 50 years (interquartile range [IQR] 48 - 52), with early menopause observed in 149 women (7%). Women were aged 20 to 45 years at the first EPDS assessment and 50-75 years at the final assessment, with reproductive age ranging from -28 to +31 years relative to the FMP. On average, women contributed 9.1 EPDS measures (SD 2.3), with a median of 10 measures (IQR 8 – 11).

Baseline characteristics are summarised in Table 1. In the primary analysis, 98% of the women were white, 55% were never smokers, 61% had high education (A-levels or higher), 86% consumed alcohol less than four times per month or not at all, and 70% reported menarche between 12 and 14 years. Participant characteristics were broadly similar in the secondary analysis, although proportions of smoking and alcohol consumption were slightly higher, and BMI was marginally greater (Table 1).

**Table 1.**
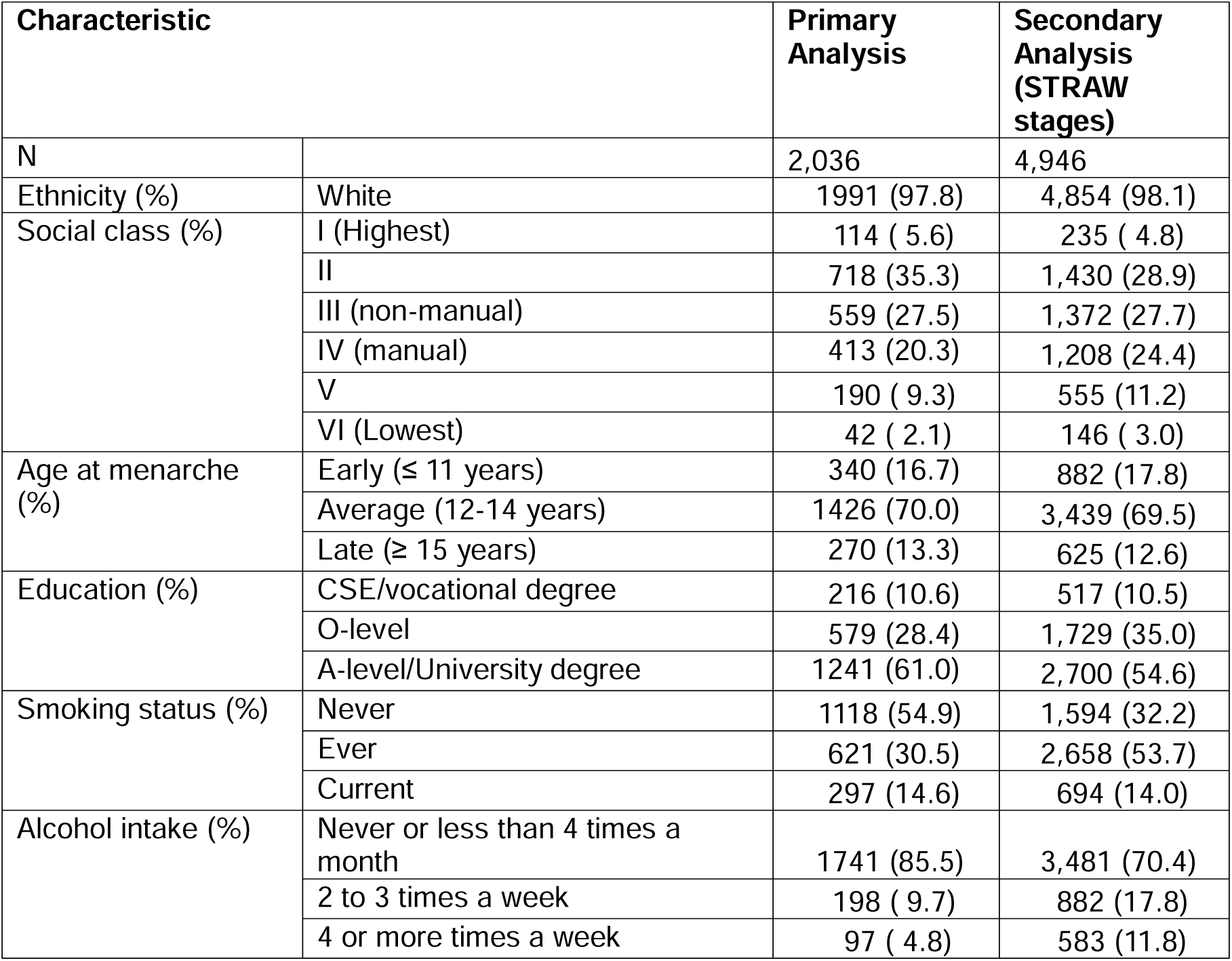
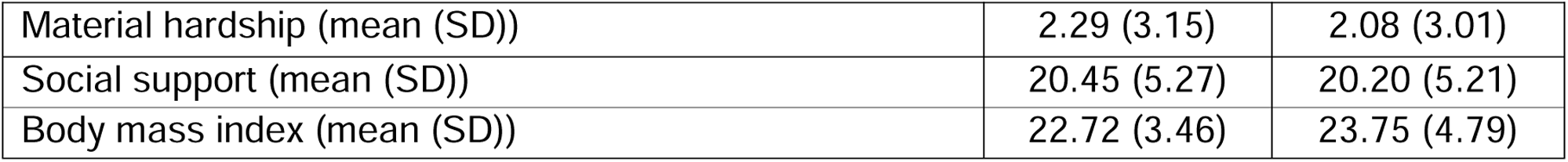
Distribution of confounders at baseline.

| Characteristic |  | Primary Analysis | Secondary Analysis (STRAW stages) |
| --- | --- | --- | --- |
| N |  | 2,036 | 4,946 |
| Ethnicity (%) | White | 1991 (97.8) | 4,854 (98.1) |
| Social class (%) | I (Highest) | 114 ( 5.6) | 235 ( 4.8) |
|  | II | 718 (35.3) | 1,430 (28.9) |
|  | III (non-manual) | 559 (27.5) | 1,372 (27.7) |
|  | IV (manual) | 413 (20.3) | 1,208 (24.4) |
|  | V | 190 ( 9.3) | 555 (11.2) |
|  | VI (Lowest) | 42 ( 2.1) | 146 ( 3.0) |
| Age at menarche (%) | Early ($\leq 11$ years) | 340 (16.7) | 882 (17.8) |
|  | Average (12-14 years) | 1426 (70.0) | 3,439 (69.5) |
| | Late ($\geq 15$ years) | 270 (13.3) | 625 (12.6) |
| Education (%) | CSE/vocational degree | 216 (10.6) | 517 (10.5) |
|  | O-level | 579 (28.4) | 1,729 (35.0) |
|  | A-level/University degree | 1241 (61.0) | 2,700 (54.6) |
| Smoking status (%) | Never | 1118 (54.9) | 1,594 (32.2) |
|  | Ever | 621 (30.5) | 2,658 (53.7) |
|  | Current | 297 (14.6) | 694 (14.0) |
| Alcohol intake (%) | Never or less than 4 times a month | 1741 (85.5) | 3,481 (70.4) |
|  | 2 to 3 times a week | 198 ( 9.7) | 882 (17.8) |
|  | 4 or more times a week | 97 ( 4.8) | 583 (11.8) |

|  |  |  |
| --- | --- | --- |
| Material hardship (mean (SD)) | 2.29 (3.15) | 2.08 (3.01) |
| Social support (mean (SD)) | 20.45 (5.27) | 20.20 (5.21) |
| Body mass index (mean (SD)) | 22.72 (3.46) | 23.75 (4.79) |

In the secondary analyses examining depressive symptoms by chronological age across menopause stages, 4,946 women contributed a total of 27,661 EPDS observations. Of these, 50% of observations occurred during the reproductive period (median age at observation 35 years), 13% during perimenopause (median age 50 years), and 37% during postmenopause (median age 59 years).

### Results for EPDS as a continuous outcome

There was no evidence of non-linear associations between reproductive age and depressive symptoms, whereas there was some evidence of non-linearity in associations between chronological age and depressive symptoms (Supplementary Table 4, Supplementary Figures 1-2). Accordingly, models included a linear term for reproductive age and both linear and quadratic terms for chronological age (See Supplementary Table 5 for full model specifications).

The trajectory of depressive symptoms across reproductive age, adjusted for chronological age is presented in Figure 2A. Depressive symptom scores showed a small increase of 0.05 points/year (95% CI 0.004 – 0.10), rising linearly from a mean EPDS score of 5.9 (95% CI 4.9 – 6.9) twenty years before the menopause to 7.4 (95% CI 6.7 – 8.0) ten years after the menopause (Supplementary Table 6). Similar results were observed when unadjusted for chronological age (0.05 points/year; 95% CI 0.04 – 0.06; Supplementary Figure 3, Supplementary Table 6).

**Figure 2.**
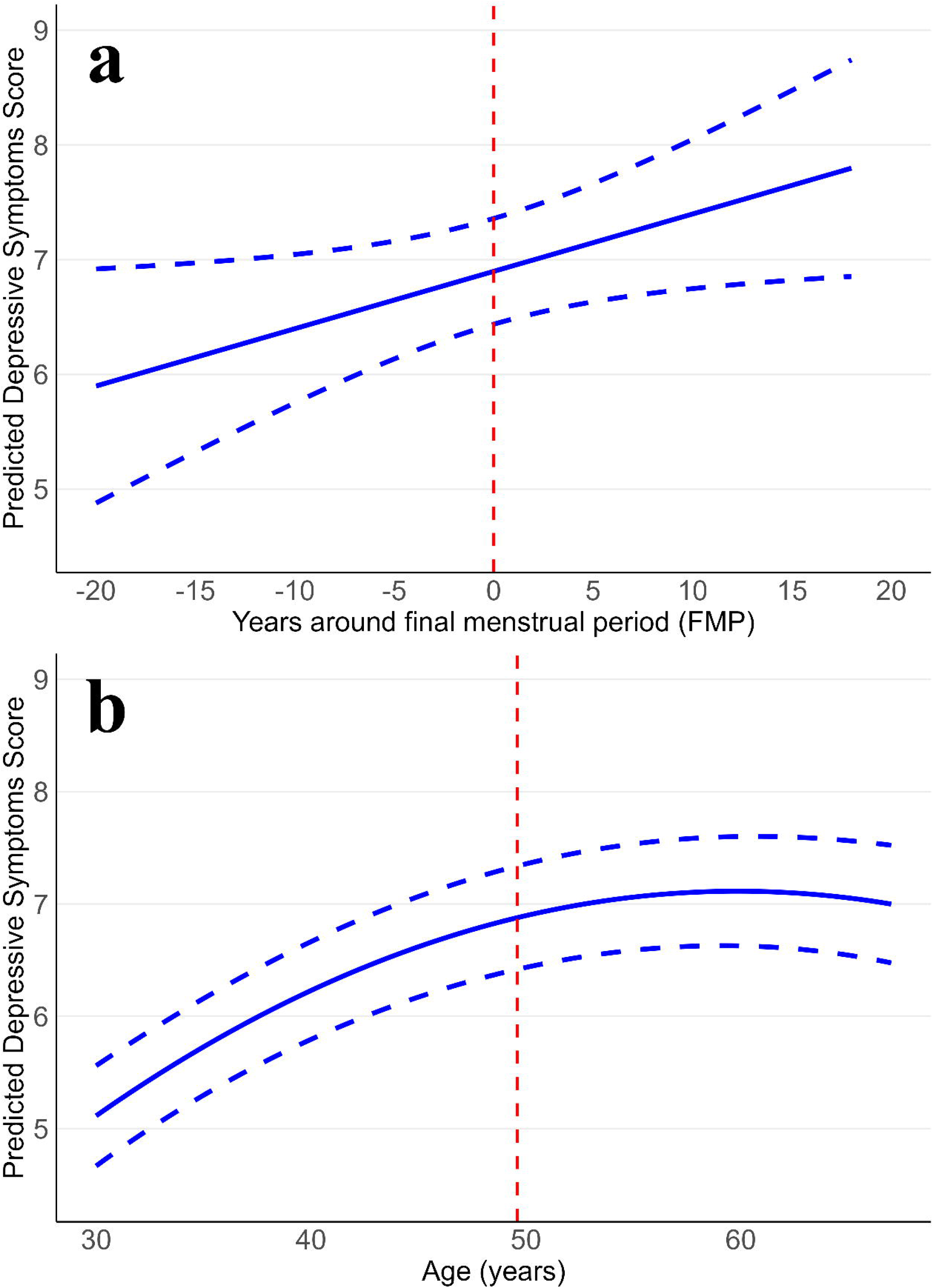
Trajectories of depressive symptoms in the years surrounding the (A) FMP and (B) chronological age (N = 2,036). Dashed vertical line corresponds to (**A**) menopause or (**B**) average age of menopause in the study.

The trajectory of depressive symptoms across chronological age is presented in Figure 2B. Depressive symptoms increased with chronological age, rising from a mean EPDS score of 5.1 (95% CI 4.7 – 5.6) at age 30 to 7.1 (95% CI 6.6 – 7.6) at age 60 (Supplementary Table 7).

Figure 3 compares the trajectories of depressive symptoms across chronological age by age at menopause (early vs. normative/late). There was little evidence that women who experienced early menopause compared to those with normative/late timing of menopause age had differing trajectories of depressive symptoms across chronological age. However, the number of women with early menopause was relatively small (n = 149) compared with the normative/late group (n = 1,887).

**Figure 3.**
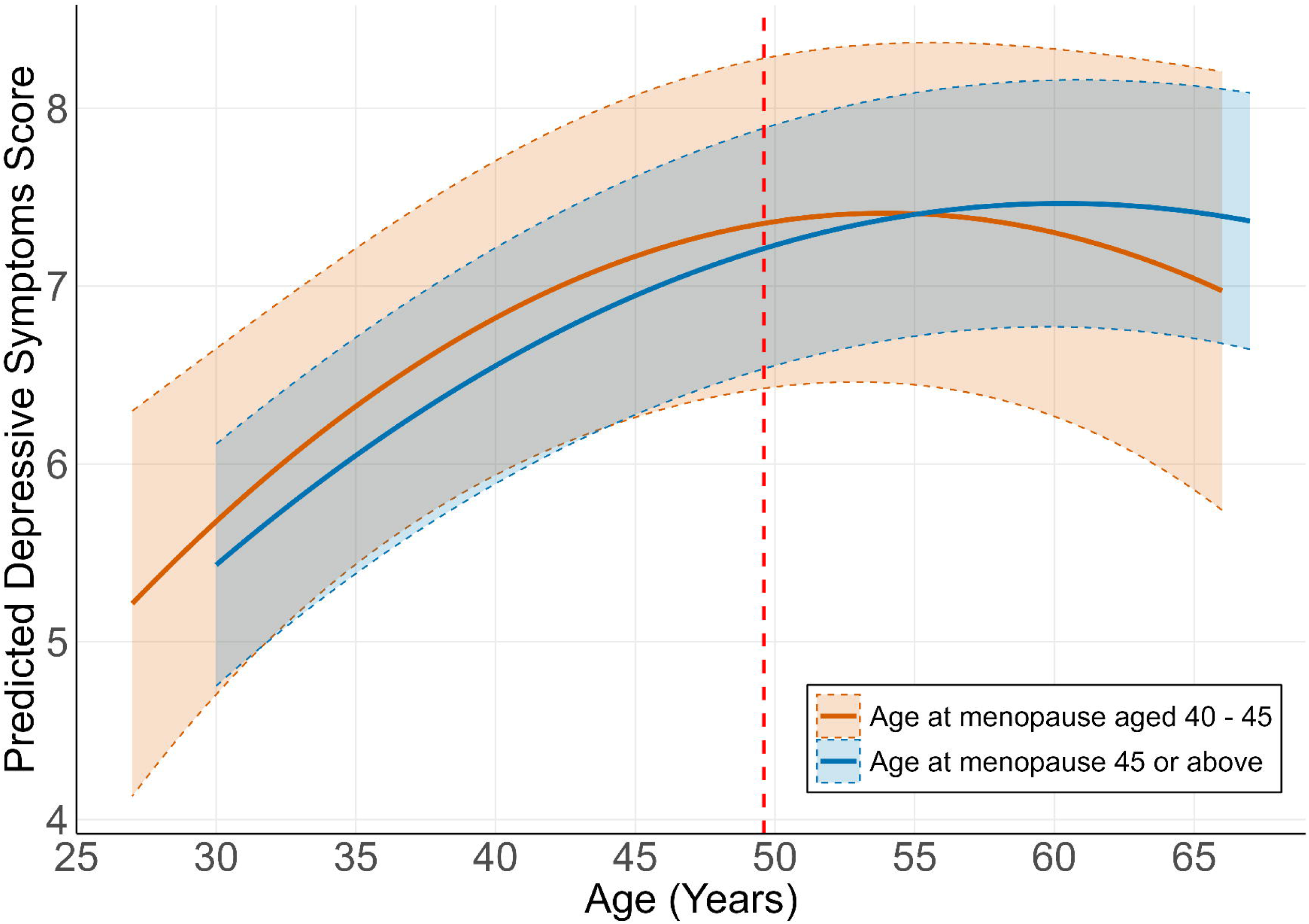
Trajectories of depressive symptoms across chronological age by age at menopause categories (N = 2,036). Dashed vertical line corresponds to average age at menopause in the study

Figure 4 shows trajectories of depressive symptoms across chronological age by menopause stages. Depressive symptoms increased with age in both the reproductive period (0.09 points per year, 95% CI 0.07 – 0.10) and perimenopause (0.07 points per year, 95% CI 0.03 – 0.12). In contrast, symptoms showed little evidence of change in the postmenopause (−0.03 points per year; 95% CI -0.06 – 0.008).

**Figure 4.**
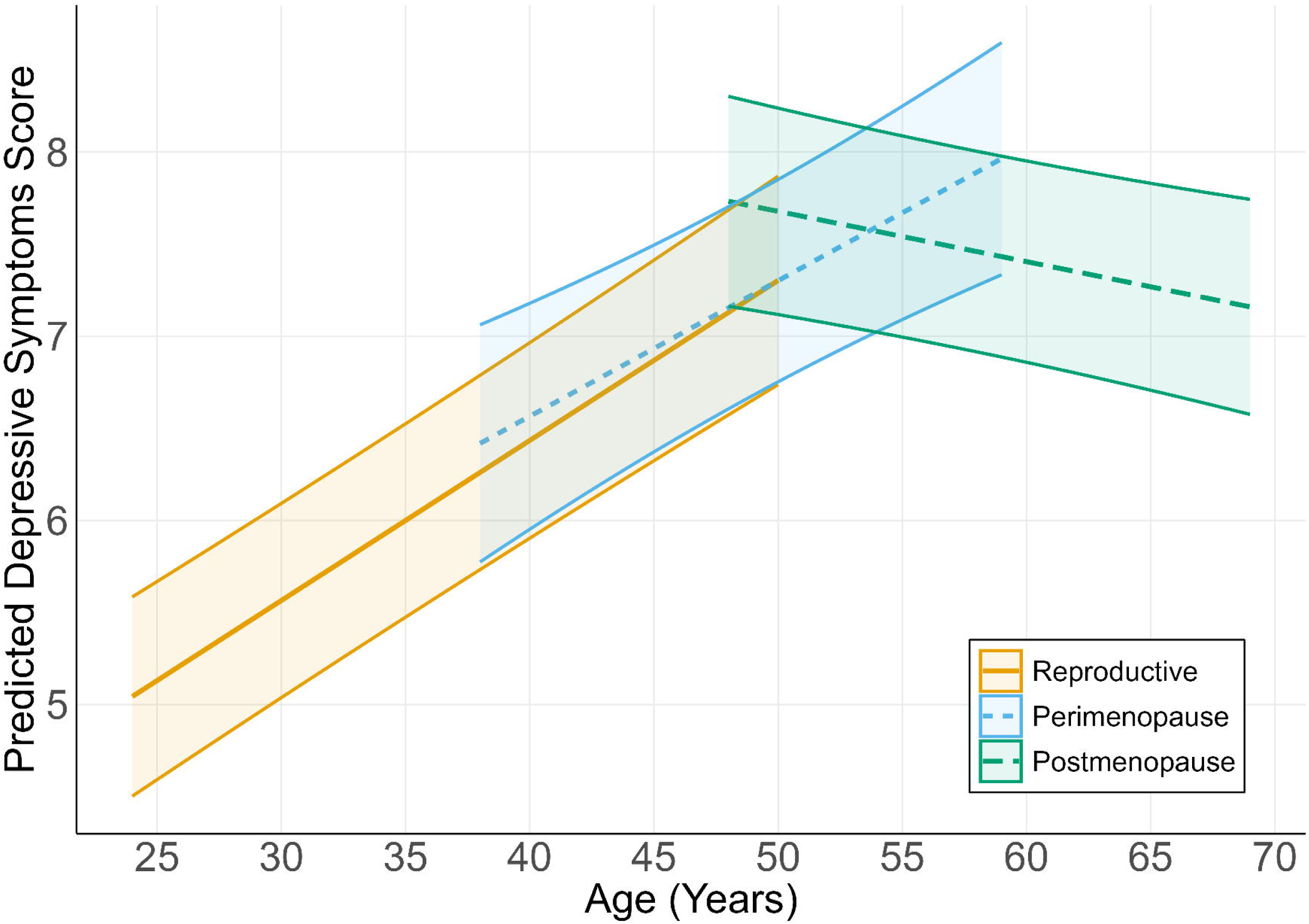
Trajectories of depressive symptoms across chronological age by menopausal stage (reproductive, perimenopause and postmenopause) (N = 4,946).

### Results for EPDS as a binary outcome

Trajectories of depressive symptoms using a binary EPDS outcome were consistent with those observed using continuous scores (Figure 5, Supplementary Figures 4 – 5). Across reproductive age, the predicted probability of depression increased modestly, rising from 0.10 (95% CI 0.06 – 0.16) twenty years before the menopause to 0.18 (95% CI 0.13 – 0.23) 10 years after the menopause (Figure 5A). This corresponds to an estimated 2% increase in the odds of depression for each one-year increase in reproductive age (OR 1.02; 95 CI 1.00 – 1.05).By contrast, across chronological age, the predicted probability of depression increased more steeply before age 50 and plateaued thereafter, rising from 0.07 (95% CI 0.06 – 0.09) at age 30 to 0.15 (95% CI 0.12 – 0.18) at age 50, and 0.16 (95% CI 0.12 –0.19) at age 60 (Figure 5B).

**Figure 5.**
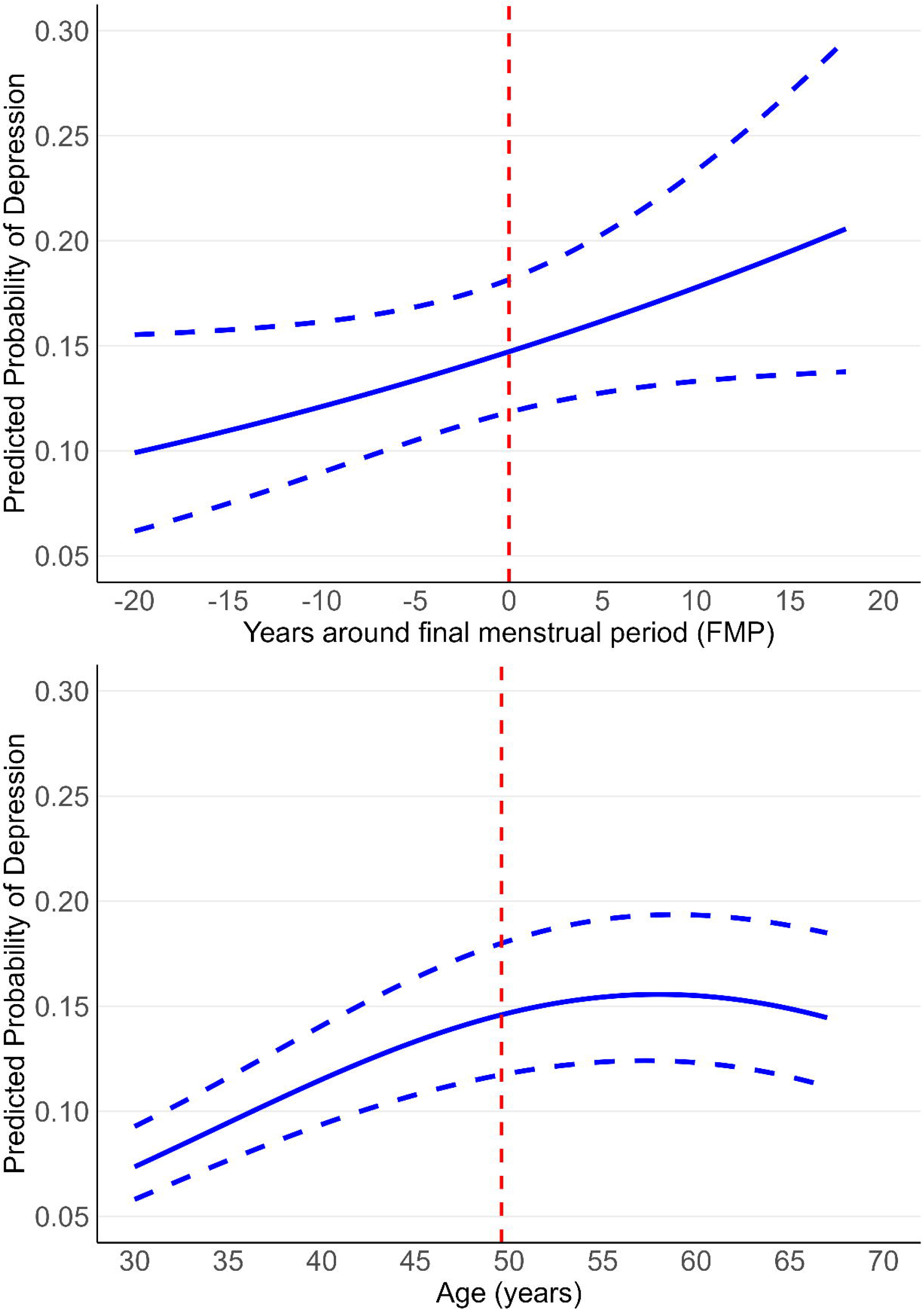
Predicted probability of depression in the years surrounding the (A) FMP and (B) chronological age (N = 2,036). Dashed vertical line corresponds to (**A**) menopause or (**B**) average age of menopause in the study.

Compared with the reproductive period, the perimenopausal period was associated with higher odds of depression (OR 1.19; 95% CI 1.03 – 1.39), whereas there was little evidence of difference in odds during postmenopause (OR 0.99; 95% CI 0.83 – 1.19) (Table 2).

**Table 2.** Random effects logistic regression of depression (EPDS ≥ 13), N = 4,946.

| Menopausal status | Odds ratio | 95% C.I. | p-value |
| --- | --- | --- | --- |
| Reproductive (ref) | 1.00 (ref) | - | - |
| Perimenopause | 1.19 | 1.03 – 1.39 | 0.02 |
| Postmenopause | 0.99 | 0.83 – 1.19 | 0.95 |

Model adjusted for age, ethnicity, social class, education, age at menarche, material hardship, social support, smoking status, body mass index and alcohol intake.

### Sensitivity analyses

Results for reproductive and chronological age models were similar when data were restricted at the 5^th^ and 95^th^ centiles of reproductive age (reproductive age range: -18 to +16 years; Supplementary Figure 6). Log-transformation of EPDS scores produced a comparable upward trend, though at a lower overall level (Supplementary Figure 7), however model residuals indicated comparable model fit. Findings were also unchanged when history of depression was included as a confounder (N = 2,014, 18,278 observations)(Supplementary Figures 8 - 9, Supplementary Table 8), suggesting that results are unlikely to be driven by pre-existing depressive symptoms.

When depressive symptoms were modelled by chronological age in the full sample of women (N = 9,277 women contributing 62,583 EPDS observations; Supplementary Figure 10), results were broadly consistent with the sample of women with an age at menopause estimate (Figure 2B). In the full sample, depressive symptoms increased with chronological age, rising from a mean EPDS score of 5.5 (95% CI 5.3 – 5.7) at age 30 to 7.6 (95% CI 7.3 –7.9) at age 70 compared with 5.1 (95% CI 4.7 – 5.6) at age 30 to 6.9 (95% CI 6.3 – 7.4) at age 70.

Ever-use of HRT was reported by 25% of women. Trajectories of depressive symptoms across reproductive age by HRT use (ever vs. never) are shown in Supplementary Figure 11. Although women who never used HRT appeared to have lower baseline levels of depressive symptoms than ever-users, confidence intervals were wide and overlapped. In both groups, depressive symptoms increased with reproductive age, with similar rates of change for ever-users (0.04 points/year; 95% CI -0.009 - 0.08) and never-users (0.04 points/year; 95% -0.02 - 0.10). The interaction between reproductive age and HRT use indicated no evidence that trajectories of depressive symptoms across reproductive age differed by HRT use (*P* = 0.84).

## Discussion

### Principal findings

In this UK community-based cohort of more than 2,000 women followed for over three decades, we examined trajectories of depressive symptoms across both reproductive and chronological age. Depressive symptoms increased modestly across midlife, however, we found little evidence of a marked increase in symptoms specifically around the menopause transition. When examined by menopause stage, symptoms increased gradually and at similar rates across the reproductive and perimenopausal periods before remaining relatively constant in the postmenopause. In complementary analyses, perimenopause was associated with higher odds of depression compared with the reproductive stage, whereas no increased odds were observed in postmenopause.

Taken together, these findings suggest that reproductive ageing appears to contribute relatively little to long-term changes in depressive symptoms beyond the broader effects of chronological ageing at the population level. However, the increased odds of depression observed during perimenopause suggest that a subgroup of women may be particularly vulnerable to depressive symptoms during this time. Importantly, any elevation in risk appears to be transient and does not persist into postmenopause.

### Strengths and Limitations

A major strength of this study is the long follow-up, with up to 11 repeated assessments of depressive symptoms over 30 years, allowing characterisation of trajectories before, during and after the menopausal transition. Age at menopause was derived from repeated prospective reports of menstrual bleeding, instead of retrospective reports, which are prone to recall error (27, 28). The availability of a wide range of relevant confounders (baseline and time-varying) is another strength.

Several limitations should be considered. EPDS assessments were less frequent during perimenopause, reducing power to detect short-term changes within a potentially brief and heterogeneous period (median ∼4 years, though estimates vary widely with reported ranges from 2 to 8 years or longer(29–31)). Short-lived increases in depressive symptoms may therefore have been difficult to capture in trajectory models estimating average symptom levels, which may help explain why elevated odds of depression were observed in stage-based analyses while changes in mean symptom levels were modest.

The main analyses were limited to women with an estimated age at menopause, resulting in data loss and potential selection bias, as included women were more socioeconomically advantaged and healthier than those excluded (Supplementary Table 9). HRT was measured crudely and could not be modelled as a time-varying variable. As HRT is typically prescribed in response to menopausal symptoms, it may act as an effect modifier rather than a confounder, but we were unable to examine this due to limited information on timing and duration of HRT use. Strong collinearity between chronological and reproductive age (r = 0.95) made it difficult to fully disentangle their independent effects. We opted not to impute missing confounder data due to the complexity of imputing multilevel longitudinal data. Missingness in confounders was generally low (Supplementary Table 10), but bias due to data not missing at random cannot be ruled out. Finally, the cohort predominantly comprised White European, parous women, limiting generalisability to more diverse populations.

### Comparison with other studies

Most previous longitudinal studies have compared depressive symptoms across menopause stages(10, 32–34). A recent meta-analysis reporting elevated odds of depression confined to perimenopause, with no increased risk observed in postmenopause relative to the reproductive period(10), consistent with our findings. However, these studies have focused on differences between menopause stages rather than examining how depressive symptoms change over time across the menopause transition. By modelling trajectories across both reproductive and chronological age, our study was able to assess whether depressive symptoms change more rapidly around the menopause transition itself.

Our findings are also consistent with latent class analyses from a large population-based cohort study showing that most women experience stable low levels of depressive symptoms across the menopause transition, while only a minority experience persistently high or increasing symptoms(35). This life-course approach helps to place findings from stage-based analyses within the broader context of midlife changes in depressive symptoms.

### Possible mechanisms

The menopause transition has been linked to depression through several biological and psychosocial mechanisms. Fluctuations in ovarian hormones, particularly oestradiol, have been proposed as mechanisms linking reproductive ageing to mood disturbances(32, 36–38), and longer lifetime exposure to endogenous oestrogens has been associated with lower risk of postmenopausal depression(11). Qualitative research suggests that menopause is often experienced within broader social and cultural contexts that emphasise ageing, loss of fertility, and changing identity(39) which may also negatively affect psychological wellbeing. However, the menopause transition coincides with midlife, a period often characterised by increased caring responsibilities, work and financial pressures, changing relationships, and emerging health concerns(40–45). Given that our trajectory analyses showed only modest increases in depressive symptoms across midlife, with little evidence of a marked change specifically around the menopause transition, our findings suggest that broader midlife and ageing-related factors may be more important drivers of population-level changes in depressive symptoms.

### Future research

Future research should explore whether there are distinct subgroups of women at higher risk of depression during the menopausal transition, and if so, clarify the underlying mechanisms driving this increased risk. Such research could help to inform more targeted prevention and treatment strategies for depression in women during perimenopause. Prior studies suggest that factors such as vasomotor symptoms(7, 34), sleep disturbance(32), and stressful life events(34) are linked to higher depressive symptoms during perimenopause. It is also possible that some women are particularly sensitive to hormonal fluctuations, such as those occurring during perimenopause, increasing vulnerability to both menopausal symptoms and depression.(46)

Clarification of the role of HRT in perimenopausal and postmenopausal depression is also needed, particularly given increasing prescribing rates over the past decade(47, 48). While HRT is effective in relieving menopausal symptoms such as hot flushes, night sweats and sleep problems(24, 49–54), it is not currently licensed to treat menopausal depression or anxiety. Clinical guidelines (including NICE, IMS, the North American Menopause Society, and the British Menopause Society) recommend that HRT can be considered for the treatment of menopausal mood symptoms(24, 52–54), but randomised clinical trial data supporting its use in this context remain limited(55–60). More precise measurements of reproductive ageing - through more frequent and detailed assessment of menstrual cycle patterns – would improve classification of menopause stage and strengthen future research examining mental health and other health outcomes across the menopause transition.

## Conclusion

We found little evidence that mean levels of depressive symptoms meaningfully change across reproductive age beyond that of chronological ageing. Although the perimenopause may represent a period of increased risk for depression, any elevation in risk appears to be transient and does not seem to persist into the postmenopause. These findings suggest that the menopause transition is unlikely to be a major driver of population-level changes in depressive symptoms, but may represent a window of vulnerability for a subgroup of women. Further research is needed to identifying women most at risk during the menopause transition to inform targeted prevention and support strategies.

## Data Availability

The informed consent obtained from ALSPAC (Avon Longitudinal Study of Parents and Children) participants does not allow the data to be made available through any third party maintained public repository. Supporting data are available from ALSPAC on request under the approved proposal number, B4425. Full instructions for applying for data access can be found here: http://www.bristol.ac.uk/alspac/researchers/access/. The ALSPAC study website contains details of all available data (http://www.bristol.ac.uk/alspac/researchers/our-data/).
Any questions regarding data or sample access should be directed to (data) or (samples).
All code used to run analyses are available on GitHub: RochelleKnight/Trajectories-of-Depressive-Symptoms-Across-the-Menopause-Transition
Archived software available from: https://doi.org/10.5281/zenodo.19388441

## List of abbreviations

STRAW: Stages of Reproductive Aging Workshop
EPDS: Edinburgh Postnatal Depressio Score
FMP: Final Menstrual Period
RCS: Restricted Cubic Spline
AIC: Akaike Information Criterion
BIC: Bayesian Information Criterion
HRT: Hormone Replacement Therapy

## Declarations

### Ethical approval and consent

Ethical approval for the study was obtained from the ALSPAC Ethics and Law Committee and the Local Research Ethics Committees. Informed consent for the use of all data collected was obtained from participants following the recommendations of the ALSPAC Ethics and Law Committee at the time. Study participation is voluntary and during all data collection sweeps, information was provided on the intended use of data. Participants can contact the study team at any time to retrospectively withdraw consent for their data to be used. The completion of a questionnaire, either on paper or online, was considered to be written consent from participants to use their data for research purposes. Full details of the approvals are available from the <u>study website</u>.

### Availability of Data and Materials

The informed consent obtained from ALSPAC (Avon Longitudinal Study of Parents and Children) participants does not allow the data to be made available through any third party maintained public repository. Supporting data are available from ALSPAC on request under the approved proposal number, B4425. Full instructions for applying for data access can be found here: http://www.bristol.ac.uk/alspac/researchers/access/. The ALSPAC study website contains details of all available data (http://www.bristol.ac.uk/alspac/researchers/our-data/).

Any questions regarding data or sample access should be directed to (data) or (samples).

All code used to run analyses are available on GitHub: RochelleKnight/Trajectories-of-Depressive-Symptoms-Across-the-Menopause-Transition Archived software available from: https://doi.org/10.5281/zenodo.19388441

## Funding

RK is supported by the Wellcome trust [228278/Z/23/Z, 218495/Z/19/Z].

AGS is supported by STAGE that has received funding from the European Union’s Horizon Europe Research and Innovation Programme under grant agreement n° 101137146 (via UKRI grant number 10099041). AF and AGS work in a Unit that is funded by the UK Medical Research Council (MC_UU_00011/1&6) and the University of Bristol.

The UK Medical Research Council and Wellcome [217065/Z/19/Z] and the University of Bristol provide core support for ALSPAC. This publication is the work of the authors and Rochelle Knight will serve as guarantor for the contents of this paper. This research was funded in whole, or in part, by the Wellcome Trust [228278/Z/23/Z, 218495/Z/19/Z]. For the purpose of Open Access, the author has applied a CC BY public copyright licence to any Author Accepted Manuscript version arising from this submission. A comprehensive list of grants funding is available on the ALSPAC website: http://www.bristol.ac.uk/alspac/external/documents/grant-acknowledgements.pdf. This research was specifically funded by the John Templeton Foundation [61356, 61356] and the Wellcome Trust [WT092830/Z/10/Z].

*The funders had no role in study design, data collection and analysis, decision to publish, or preparation of the manuscript*.

### Author Contributions

**Rochelle Knight:** Conceptualization, Methodology, Software, Data Curation, Writing – Original Draft. **Abigail Fraser:** Conceptualization, Methodology, Writing - Review & Editing, Supervision. **Carol Joinson:** Conceptualization, Methodology, Writing - Review & Editing, Supervision. **Ana Gonçalves Soares:** Conceptualization, Methodology, Software, Writing - Review & Editing, Supervision

## Acknowledgements

We are extremely grateful to all the families who took part in this study, the midwives for their help in recruiting them, and the whole ALSPAC team, which includes data collection staff, data and administrations staff, technical managers and the technical staff with the Bristol Bioresource Laboratory, based within the University of Bristol.

## Additional files

### Additional file 1: Supplementary tables and figures

Tables S1 – S10 and Figures S1 – S11.

Table S1. Description of EPDS timepoints for women included in the main analysis. Table S2. Derivation of the EPDS depressive symptom score outcome variable from Avon Longitudinal Study of Parents and Children (ALSPAC). Table S3. Derivation of the confounder variables from Avon Longitudinal Study of Parents and Children (ALSPAC). Table S4. Comparison of model fit across linear, quadratic and restricted cubic spline models. Table S5. Overview of Multilevel Model Structures and Covariate Adjustments. All models included a participant level random intercept. Table S6. Average predicted depressive symptom scores across reproductive age. Supplementary Table S7. Average predicted depressive symptom scores across chronological age. Table S8. Association between menopause stage and depression, further adjusting for baseline history of depression as a confounder. Table S9. Comparison of confounders at baseline between women with and without an age at menopause. Table S10. Proportion of observations with missing data for each confounder.

Figure S1. Comparison of model fit for trajectories of depressive symptoms in the years surrounding the final menstrual period. Figure S2. Comparison of model fit for trajectories of depressive symptoms across chronological age. Figure S3. Trajectory of depressive symptoms in the years surrounding the final menstrual period unadjusted for chronological age. Figure S4. Predicted probability of depression across chronological age, with an interaction for menopausal stage (pre-, peri-, and post-menopause). Figure S5. Predicted probability of depression across chronological age, with an interaction for age at menopause categories. Figure S6. Trajectories of depressive symptoms in the years surrounding the final menstrual period (A) and chronological age (B), restricted at the 5th and 95th centiles of age at final menstrual period. Figure S7. Trajectories of depressive symptoms in the years surrounding the final menstrual period (A) and chronological age (B). EPDS score was log transformed prior to modelling. Figure S8. Trajectories of depressive symptoms in the years surrounding the (A) final menstrual period and (B) chronological age, further adjusting for baseline history of depression as a confounder. Figure S9. Predicted probability of depression in the years surrounding the (A) final menstrual period and (B) chronological age, further adjusting for baseline history of depression as a confounder. Figure S10. Trajectory of depressive symptoms across chronological age in the full population (N= 9,277). Figure S11. Trajectories of depressive symptoms across reproductive age by HRT use (Ever vs. Never).

### Additional file 2: Supplementary methods

Supplementary Methods on Estimation of age at natural menopause and reproductive stage.

## References

1. Bromet E, Andrade LH, Hwang I, Sampson NA, Alonso J, de Girolamo G, et al. Cross-national epidemiology of DSM-IV major depressive episode. BMC Med. 2011;9:90.

2. Kessler RC, McGonagle KA, Swartz M, Blazer DG, Nelson CB. Sex and depression in the National Comorbidity Survey. I: Lifetime prevalence, chronicity and recurrence. J Affect Disord. 1993;29(2-3):85–96.

3. Salk RH, Hyde JS, Abramson LY. Gender differences in depression in representative national samples: Meta-analyses of diagnoses and symptoms. Psychol Bull. 2017;143(8):783–822.

4. Soares CN, Zitek B. Reproductive hormone sensitivity and risk for depression across the female life cycle: a continuum of vulnerability? J Psychiatry Neurosci. 2008;33(4):331–43.

5. Arevalo MA, Azcoitia I, Garcia-Segura LM. The neuroprotective actions of oestradiol and oestrogen receptors. Nat Rev Neurosci. 2015;16(1):17–29.

6. El Khoudary SR, Greendale G, Crawford SL, Avis NE, Brooks MM, Thurston RC, et al. The menopause transition and women’s health at midlife: a progress report from the Study of Women’s Health Across the Nation (SWAN). Menopause. 2019;26(10):1213–27.

7. Avis NE, Brambilla D, McKinlay SM, Vass K. A longitudinal analysis of the association between menopause and depression. Results from the Massachusetts Women’s Health Study. Ann Epidemiol. 1994;4(3):214–20.

8. Kaufert PA, Gilbert P, Tate R. The Manitoba Project: a re-examination of the link between menopause and depression. Maturitas. 2008;61(1-2):54–66.

9. Jones HJ, Minarik PA, Gilliss CL, Lee KA. Depressive symptoms associated with physical health problems in midlife women: A longitudinal study. J Affect Disord. 2020;263:301–9.

10. Badawy Y, Spector A, Li Z, Desai R. The risk of depression in the menopausal stages: A systematic review and meta-analysis. J Affect Disord. 2024;357:126–33.

11. Georgakis MK, Thomopoulos TP, Diamantaras AA, Kalogirou EI, Skalkidou A, Daskalopoulou SS, et al. Association of Age at Menopause and Duration of Reproductive Period With Depression After Menopause: A Systematic Review and Meta-analysis. JAMA Psychiatry. 2016;73(2):139–49.

12. Marsh WK, Bromberger JT, Crawford SL, Leung K, Kravitz HM, Randolph JF, et al. Lifelong estradiol exposure and risk of depressive symptoms during the transition to menopause and postmenopause. Menopause. 2017;24(12):1351–9.

13. Harris PA, Taylor R, Thielke R, Payne J, Gonzalez N, Conde JG. Research electronic data capture (REDCap)--a metadata-driven methodology and workflow process for providing translational research informatics support. J Biomed Inform. 2009;42(2):377–81.

14. Boyd A, Golding J, Macleod J, Lawlor DA, Fraser A, Henderson J, et al. Cohort Profile: the ‘children of the 90s’--the index offspring of the Avon Longitudinal Study of Parents and Children. Int J Epidemiol. 2013;42(1):111–27.

15. Fraser A, Macdonald-Wallis C, Tilling K, Boyd A, Golding J, Davey Smith G, et al. Cohort Profile: the Avon Longitudinal Study of Parents and Children: ALSPAC mothers cohort. Int J Epidemiol. 2013;42(1):97–110.

16. Major-Smith D, Heron J, Fraser A, Lawlor DA, Golding J, Northstone K. The Avon Longitudinal Study of Parents and Children (ALSPAC): a 2022 update on the enrolled sample of mothers and the associated baseline data. Wellcome Open Res. 2022;7:283.

17. Northstone K, Lewcock M, Groom A, Boyd A, Macleod J, Timpson N, et al. The Avon Longitudinal Study of Parents and Children (ALSPAC): an update on the enrolled sample of index children in 2019. Wellcome Open Res. 2019;4:51.

18. Harlow SD, Gass M, Hall JE, Lobo R, Maki P, Rebar RW, et al. Executive summary of the Stages of Reproductive Aging Workshop + 10. Menopause. 2012;19(4):387–95.

19. Cox JL, Holden JM, Sagovsky R. Detection of postnatal depression. Development of the 10-item Edinburgh Postnatal Depression Scale. Br J Psychiatry. 1987;150:782–6.

20. Murray L, Carothers AD. The validation of the Edinburgh Post-natal Depression Scale on a community sample. Br J Psychiatry. 1990;157:288–90.

21. Knight R, Fraser A, Joinson C, Soares A. Estimating age of menopause in mothers in the ALSPAC Study: A data note [version 1; peer review: awaiting peer review]. Wellcome Open Research. 2025;10(631).

22. An S, Ren S, Ma J, Zhang Y. Association of Depression with Age at Natural Menopause: A Cross-Sectional Analysis with NHANES Data. Int J Womens Health. 2025;17:211–20.

23. Harlow BL, Wise LA, Otto MW, Soares CN, Cohen LS. Depression and Its Influence on Reproductive Endocrine and Menstrual Cycle Markers Associated With Perimenopause: The Harvard Study of Moods and Cycles. Archives of General Psychiatry. 2003;60(1):29–36.

24. NICE guideline NG23. Menopause: identification and management 2024 [Available from: https://www.nice.org.uk/guidance/NG23.

25. Ossewaarde ME, Bots ML, Verbeek ALM, Peeters PHM, van der Graaf Y, Grobbee DE, et al. Age at Menopause, Cause-Specific Mortality and Total Life Expectancy. Epidemiology. 2005;16(4):556–62.

26. Bromberger JT, Kravitz HM. Mood and menopause: findings from the Study of Women’s Health Across the Nation (SWAN) over 10 years. Obstet Gynecol Clin North Am. 2011;38(3):609–25.

27. Colditz GA, Stampfer MJ, Willett WC, Stason WB, Rosner B, Hennekens CH, et al. REPRODUCIBILITY AND VALIDITY OF SELF-REPORTED MENOPAUSAL STATUS IN A PROSPECTIVE COHORT STUDY. American Journal of Epidemiology. 1987;126(2):319–25.

28. Hahn RA, Eaker E, Rolka H. Reliability of reported age at menopause. Am J Epidemiol. 1997;146(9):771–5.

29. Delamater L, Santoro N. Management of the Perimenopause. Clin Obstet Gynecol. 2018;61(3):419–32.

30. Burger H, Woods NF, Dennerstein L, Alexander JL, Kotz K, Richardson G. Nomenclature and endocrinology of menopause and perimenopause. Expert Rev Neurother. 2007;7(11 Suppl):S35–43.

31. Wegrzynowicz AK, Walls AC, Godfrey M, Beckley A. Insights into Perimenopause: A Survey of Perceptions, Opinions on Treatment, and Potential Approaches. Women (Basel). 2025;5(1).

32. Freeman EW, Sammel MD, Liu L, Gracia CR, Nelson DB, Hollander L. Hormones and Menopausal Status as Predictors of Depression in Women in Transition to Menopause. Archives of General Psychiatry. 2004;61(1):62.

33. Bromberger JT, Matthews KA, Schott LL, Brockwell S, Avis NE, Kravitz HM, et al. Depressive symptoms during the menopausal transition: The Study of Women’s Health Across the Nation (SWAN). Journal of Affective Disorders. 2007;103(1-3):267–72.

34. Cohen LS, Soares CN, Vitonis AF, Otto MW, Harlow BL. Risk for New Onset of Depression During the Menopausal Transition. Archives of General Psychiatry. 2006;63(4):385.

35. Hickey M, Schoenaker DA, Joffe H, Mishra GD. Depressive symptoms across the menopause transition: findings from a large population-based cohort study. Menopause. 2016;23(12):1287–93.

36. Freeman EW. Associations of depression with the transition to menopause. Menopause. 2010;17(4):823–7.

37. Schmidt PJ, Rubinow DR. Sex hormones and mood in the perimenopause. Ann N Y Acad Sci. 2009;1179:70–85.

38. Bromberger JT, Schott LL, Kravitz HM, Sowers M, Avis NE, Gold EB, et al. Longitudinal Change in Reproductive Hormones and Depressive Symptoms Across the Menopausal Transition. Archives of General Psychiatry. 2010;67(6):598.

39. De Salis I, Owen-Smith A, Donovan JL, Lawlor DA. Experiencing menopause in the UK: The interrelated narratives of normality, distress, and transformation. Journal of Women & Aging. 2018;30(6):520–40.

40. Wood K, McCarthy S, Pitt H, Randle M, Thomas SL. Women’s experiences and expectations during the menopause transition: a systematic qualitative narrative review. Health Promotion International. 2025;40(1).

41. Dennerstein L, Lehert P, Burger H, Guthrie J. Sexuality. The American Journal of Medicine. 2005;118(12, Supplement 2):59–63.

42. Gyllstrom ME, Schreiner PJ, Harlow BL. Perimenopause and depression: strength of association, causal mechanisms and treatment recommendations. Best Pract Res Clin Obstet Gynaecol. 2007;21(2):275–92.

43. Manderson L. The social and cultural context of sexual function among middle-aged women. Menopause. 2005;12(4):361–2.

44. Soares CN. Depression during the menopausal transition: window of vulnerability or continuum of risk? Menopause. 2008;15(2):207–9.

45. Bromberger JT, Schott L, Kravitz HM, Joffe H. Risk factors for major depression during midlife among a community sample of women with and without prior major depression: are they the same or different? Psychological Medicine. 2015;45(8):1653–64.

46. Bromberger JT, Epperson CN. Depression During and After the Perimenopause: Impact of Hormones, Genetics, and Environmental Determinants of Disease. Obstet Gynecol Clin North Am. 2018;45(4):663–78.

47. Burkard T, Moser M, Rauch M, Jick SS, Meier CR. Utilization pattern of hormone therapy in UK general practice between 1996 and 2015: a descriptive study. Menopause. 2019;26(7):741–9.

48. Iversen L, Delaney EK, Hannaford PC, Black C. Menopause-related workload in general practice 1996–2005: a retrospective study in the UK. Family Practice. 2010;27(5):499–506.

49. Levy B, Simon JA. A Contemporary View of Menopausal Hormone Therapy. Obstet Gynecol. 2024;144(1):12–23.

50. Davis SR. Use of Testosterone in Postmenopausal Women. Endocrinol Metab Clin North Am. 2021;50(1):113–24.

51. Piette PCM. The pharmacodynamics and safety of progesterone. Best Pract Res Clin Obstet Gynaecol. 2020;69:13–29.

52. Baber RJ, Panay N, Fenton A. 2016 IMS Recommendations on women’s midlife health and menopause hormone therapy. Climacteric. 2016;19(2):109-50.

53. The 2022 hormone therapy position statement of The North American Menopause Society. Menopause. 2022;29(7):767–94.

54. Hamoda H, Panay N, Pedder H, Arya R, Savvas M. The British Menopause Society & Women’s Health Concern 2020 recommendations on hormone replacement therapy in menopausal women. Post Reprod Health. 2020;26(4):181–209.

55. Toffol E, Heikinheimo O, Partonen T. Hormone therapy and mood in perimenopausal and postmenopausal women: a narrative review. Menopause. 2015;22(5):564–78.

56. Soares CN, Arsenio H, Joffe H, Bankier B, Cassano P, Petrillo LF, et al. Escitalopram versus ethinyl estradiol and norethindrone acetate for symptomatic peri- and postmenopausal women: impact on depression, vasomotor symptoms, sleep, and quality of life. Menopause. 2006;13(5):780–6.

57. Gleason CE, Dowling NM, Wharton W, Manson JE, Miller VM, Atwood CS, et al. Effects of Hormone Therapy on Cognition and Mood in Recently Postmenopausal Women: Findings from the Randomized, Controlled KEEPS-Cognitive and Affective Study. PLoS Med. 2015;12(6):e1001833; discussion e.

58. Joffe H, Petrillo LF, Koukopoulos A, Viguera AC, Hirschberg A, Nonacs R, et al. Increased estradiol and improved sleep, but not hot flashes, predict enhanced mood during the menopausal transition. J Clin Endocrinol Metab. 2011;96(7):E1044–54.

59. Schmidt PJ, Nieman L, Danaceau MA, Tobin MB, Roca CA, Murphy JH, et al. Estrogen replacement in perimenopause-related depression: A preliminary report. American Journal of Obstetrics and Gynecology. 2000;183(2):414–20.

60. Gordon JL, Rubinow DR, Eisenlohr-Moul TA, Xia K, Schmidt PJ, Girdler SS. Efficacy of Transdermal Estradiol and Micronized Progesterone in the Prevention of Depressive Symptoms in the Menopause Transition: A Randomized Clinical Trial. JAMA Psychiatry. 2018;75(2):149–57.

