## Supplementary Methods for "Trajectories of Depressive Symptoms Across the Menopause Transition: A Longitudinal Analysis from a Prospective UK Cohort Study"

### Estimation of age at natural menopause and reproductive stage

Age at natural menopause was estimated using self-reported menstrual bleeding data, as described in detail elsewhere^[1]^. Briefly, women provided information on their menstrual cycles at up to eight questionnaire timepoints. At each timepoint they reported:

i. The date of their last menstrual period (LMP) (can be reported incompletely);
ii. Whether they had experienced a menstrual period in the previous 3 months (yes/no);
iii. Whether they had experienced a menstrual period in the previous 12 months (yes/no).

Where possible, the date of the LMP was estimated for each timepoint. The final menstrual period (FMP) was defined as the last reported menstrual period followed by at least 12 consecutive months of amenorrhea. Age at menopause was defined as the age at the FMP.

#### Assignment of reproductive stage

At each of the eight timepoints, women were classified into one of three mutually exclusive reproductive stages based on menstrual bleeding patterns, in accordance with the STRAW^[2]^ criteria:

i. Reproductive (STRAW category -4, -3b and -3a)
ii. Perimenopause (STRAW category -2, -1 and +1a)
iii. Postmenopause (STRAW category ≥ +1b, irrespective of the years since menopause))

This approach allowed inclusion of women without an estimated ANM. For example, women who could not recall the date of their last menstrual period but reported no bleeding in the preceding 12 months could be classified as postmenopausal. Similarly, women who had not yet reached menopause by the final follow-up could still be classified as perimenopausal based on bleeding patterns.

STRAW staging was applied at each timepoint, with consistency checks to ensure biologically plausible progression through reproductive stages. Because reproductive stage was determined using menstrual bleeding patterns, factors known to influence menstruation were considered, including surgery to reproductive organs, use of hormonal contraception or hormone replacement therapy (HRT), and other causes such as chemotherapy, radiotherapy, endometrial ablation, pregnancy, or breastfeeding. Full details of censoring and exclusions are reported elsewhere^[1]^.

#### Definition of recent menstrual activity

To distinguish the reproductive from the perimenopausal period, menstrual activity was classified as:

- **Recent period:** LMP within the last 60 days
- **No recent period:** LMP is ≥ 60 days and ≤ 365 days ago

Information on cycle regularity was also collected. Please see below.

| **ALSPAC Variable** | **Question(s)/measure(s)** | **Responses** | **Final variable coding** |
| --- | --- | --- | --- |
| **Period regularity** |  |  |  |
| fm1ob130, fm2ob130, fm3ob130, fm4ob130 | Are your periods regular? | 1: Yes, every 28-30 days  2: Yes, < every 28 days  3: Yes, every 30 days  4: No | Recode (4=0) (1-3 = 1)  0: Irregular periods  1: Regular periods |
| t4837, V4837 | Please describe your most recent periods:  Are your periods irregular? | 1: Very  2: Moderately  3: Mildly  4: Not at all | Recode (1/2 = 0) (3/4=0)  0: Irregular periods  1: Regular periods |
| U1050 | Are your periods regular? | 1: Yes, every 23 days or less  2: Yes, every 24-35 days  3: Yes, > every 35 days  4: No | Recode (1-3 = 1) (4=0)  0: Irregular periods  1: Regular periods |

#### Assigning the reproductive and perimenopausal period

Menstrual activity and cycle regularity were jointly used to distinguish the reproductive from the perimenopausal period at each timepoint as shown below.

| **Period regularity** | **Recent period** | **No recent period** |
| --- | --- | --- |
| **Regular** | Reproductive | Perimenopause |
| **NA** | NA | Perimenopause |
| **Irregular** | Perimenopause | Perimenopause |

Where ANM was available, perimenopause was additionally defined as the period spanning from three years prior to ANM up to one year after ANM. Within the three years prior to ANM, any timepoints previously assigned as the reproductive period were retained.

#### Assigning the postmenopausal period

Where ANM was available, timepoints that were >1 year post-ANM were assigned as postmenopausal. Where ANM was not available, women reporting no menstrual period in the preceding 12 months (yes/no response options) were classified as postmenopausal at that timepoint.

#### Additional data on menopausal status

In one questionnaire (Questionnaire MB), reproductive stage status was assessed using a different set of questions. Women were asked:

- Which of the following statements best describes your current
- menopause status?:
  1. I have been through the menopause
  2. I am going through the menopause now
  3. I have not yet started going through the menopause
  4. I am not sure
- When their last menstrual period had occurred:
  1. Within the past 3 months
  2. 4-12 months ago
  3. More than 12 months ago

Women were not assigned a reproductive stage if they reported:

- Their period had stopped as they had a hysterectomy
- Their period had stopped for other or unknown reasons
- They were pregnant or breastfeeding

|  | **Period in last 3 months** | **Period 4-12 months ago** | **Period more than 12 months ago** |
| --- | --- | --- | --- |
| **Not started going through the menopause** | Reproductive | Perimenopause | Perimenopause |
| **Going through the menopause** | Perimenopause | Perimenopause | Postmenopause |
| **Been through the menopause** | NA | NA | Postmenopause |

Reproductive stage for this questionnaire was then assigned using the classification shown below.

Women reporting the use of current contraceptives or HRT were assigned a reproductive stage only based on their response to their current menopausal status.

#### Consistency checks

The above process resulted in women having up to nine timepoints with an assigned reproductive stage. These timepoints were then checked to ensure that each woman’s reproductive status progressed in the correct chronological order over time.

**Consistency check 1: Woman is classified as reproductive after being classified as perimenopausal**

**Scenario 1.1**

A woman is classified as being in the perimenopause, then classified as reproductive at the next timepoint. If, for at least the two subsequent timepoints, she is classified as either in the perimenopause or postmenopause, her earlier classification of reproductive is changed to perimenopause.

| **Timepoint 1** | **Timepoint 2** | **Timepoint 3** | **Timepoint 4** | **Timepoint 5** |
| --- | --- | --- | --- | --- |
| Perimenopause | Reproductive | Perimenopause | Perimenopause | Postmenopause |
| Perimenopause | Reproductive | Postmenopause | Postmenopause | Postmenopause |
| Reproductive | Perimenopause | Reproductive | Perimenopause | Postmenopause |

**Scenario 1.2**

A woman is classified as being in the perimenopause for two subsequent timepoints, then classified as reproductive. The timepoint following the reproductive classification is either perimenopause, postmenopause or there are no further timepoints with STRAW classification. Her classification of reproductive is changed to perimenopause.

| **Timepoint 1** | **Timepoint 2** | **Timepoint 3** | **Timepoint 4** | **Timepoint 5** |
| --- | --- | --- | --- | --- |
| Perimenopause | Perimenopause | Reproductive | NA |  |
| Perimenopause | Perimenopause | Reproductive | Perimenopause | NA |
| Perimenopause | Perimenopause | Reproductive | Postmenopause | NA |

**Scenario 1.3**

A women is classified as being in the perimenopause, then for the following two timepoints as reproductive. The classification of perimenopause is changed to reproductive.

| **Timepoint 1** | **Timepoint 2** | **Timepoint 3** | **Timepoint 4** | **Timepoint 5** |
| --- | --- | --- | --- | --- |
| Perimenopause | Reproductive | Reproductive | Perimenopause | Perimenopause |
| Perimenopause | Reproductive | Reproductive | Reproductive | Perimenopause |
| Reproductive | Perimenopause | Reproductive | Reproductive | NA |

**Scenario 1.4**

A women is classified as being in the perimenopause, then classified as reproductive at the next timepoint. There are no further timepoints classified as reproductive. All timepoints prior to the perimenopause classification are either reproductive or the perimenopause classification was the first timepoint available with a STRAW classification. The final reproductive classification is either only followed by one further timepoint with a classification of perimenopause or postmenopause, or is the final timepoint with a STRAW classification.

| **Timepoint 1** | **Timepoint 2** | **Timepoint 3** | **Timepoint 4** | **Timepoint 5** |
| --- | --- | --- | --- | --- |
| Reproductive | Perimenopause | Reproductive | NA |  |
| Reproductive | Perimenopause | Reproductive | Perimenopause | NA |
| Reproductive | Perimenopause | Reproductive | Postmenopause | NA |
| Perimenopause | Reproductive | NA |  |  |
| Perimenopause | Reproductive | Perimenopause | NA |  |
| Perimenopause | Reproductive | Postmenopause | NA |  |

If the women’s age at the reproductive timepoint is ≤ 3 years below her age at menopause or the population mean age of menopause if ANM is unavailable, the classification of reproductive should be changed to perimenopause. If the reproductive timepoint is followed a postmenopause classification or is the final available timepoint with a straw classification, and the women’s age at the reproductive timepoint is greater than the population mean age of menopause, the classification of reproductive should be changed to postmenopause. Else the perimenopause timepoint should be changed to reproductive.

**Consistency check 2: Woman is classified as reproductive or perimenopausal after being classified as postmenopause.**

**Scenario 2.1**

A women is classified as being postmenopause, then as either reproductive or perimenopausal at the following timepoint. There are no further timepoints with a reproductive stage classification. This will **only** occur when there is no ANM available for the women and we only have information that she has not bled in the last 12 months. The woman then comes back at a later timepoint and says she has had another bleed.

If the women’s age at the reproductive timepoint is less than the population mean age of menopause, the postmenopause classification is changed to reproductive. If the women’s age at the reproductive timepoint is greater than the population mean age of menopause, the reproductive timepoint is changed to postmenopause.

If the women’s age at the perimenopause timepoint is less than the population mean age of menopause, the postmenopause classification is changed to perimenopause. If the women’s age at the perimenopause timepoint is greater than the population mean age of menopause, the reproductive timepoint is changed to postmenopause.

| **Timepoint 1** | **Timepoint 2** | **Timepoint 3** | **Age at Timepoint 3** | **Change to** |
| --- | --- | --- | --- | --- |
| Postmenopause | Reproductive | NA | 45 | Postmenopause to reproductive |
| Postmenopause | Reproductive | NA | 51 | Reproductive to postmenopause |
| Postmenopause | Perimenopause | NA | 45 | Postmenopause to perimenopause |
| Postmenopause | Perimenopause | NA | 51 | Perimenopause to postmenopause |

**Consistency check 3: Woman is classified as postmenopause prior to being classified as perimenopause or reproductive.**

**Scenario 3.1**

| **Timepoint 1** | **Timepoint 2** | **Timepoint 3** | **Change to** |
| --- | --- | --- | --- |
| Postmenopause | Reproductive | Reproductive | Reproductive |
| Postmenopause | Perimenopause | Perimenopause | Perimenopause |

A woman is classified as being postmenopause, then classified as reproductive or perimenopause for the following two timepoints. Her classification of postmenopause should be changed to reproductive or perimenopause.

**Consistency check 4: Woman switches between reproductive and perimenopause.**

**Scenario 4.1**

For all further scenarios in which a woman switches between perimenopause and reproductive and we have been unable to change inaccuracies based on any of the above, we assign reproductive stage based on age and time to menopause.

If the women’s age at the timepoint is more than 3 years below her ANM or population mean age of menopause, she is classified as reproductive. Otherwise she is classified as perimenopause.

Example:

| **Timepoint 1** | **Timepoint 2** | **Timepoint 3** | **Timepoint 4** | **Timepoint 5** | **Timepoint 6** |
| --- | --- | --- | --- | --- | --- |
| Reproductive | Perimenopause | Reproductive | Perimenopause | Reproductive | Menopause transition |

The columns highlighted in yellow are assigned menopausal status based on age at timepoint. Timepoints 1 and 6 are not changed.

#### Assigning reproductive stage to EPDS assessments

Reproductive stage was assigned at up to nine timepoints based on menstrual bleeding patterns and classified according to STRAW criteria, as described above. The EPDS was measured at 11 separate questionnaires, which did not always coincide with when the menstrual questions were asked. Therefore, reproductive stage for each EPDS assessment was assigned using information from the nearest observed reproductive stage classification.

For each women, the observed reproductive stage classifications were first used to identify:

- The latest timepoint classified as reproductive
- The earliest and latest timepoints classified as perimenopausal
- The earliest timepoint classified as postmenopausal

Where a women had only a single observed perimenopausal timepoint, a ±6-month window around that assessment was used to assign EPDS timepoints.

Using these boundaries, reproductive stage was assigned to each EPDS timepoint as follows:

- **Reproductive**: EPDS assessments occurring before the latest observed reproductive stage timepoint.
- **Perimenopausal**: EPDS assessments occurring between the first and last observed perimenopausal timepoints (or within ±6-months where only one perimenopausal observation was available).
- **Postmenopausal**: EPDS assessments occurring after the earliest observed postmenopausal timepoint.

Additionally, where ANM was available, perimenopause was defined as the period spanning from three years prior to ANM up to one year after ANM, and postmenopausal thereafter (>1 year post-ANM). Within the three years prior to ANM, any previously assigned reproductive status was retained.

1.	Knight, R., et al., *Estimating age of menopause in mothers in the ALSPAC Study: A data note [version 1; peer review: awaiting peer review].* Wellcome Open Research, 2025. **10**(631).10.12688/wellcomeopenres.24760.1

2.	Harlow, S.D., et al., *Executive summary of the Stages of Reproductive Aging Workshop + 10.* Menopause, 2012. **19**(4): p. 387-395.10.1097/gme.0b013e31824d8f40
