## Supplementary Tables and Figures for "Trajectories of Depressive Symptoms Across the Menopause Transition: A Longitudinal Analysis from a Prospective UK Cohort Study"

**Trajectories of Depressive Symptoms Across the Menopause Transition: A Longitudinal Analysis From the Avon Longitudinal Study of Parents and Children (ALSPAC)**

Rochelle Knight, Carol Joinson, Abigail Fraser, Ana Gonçalves Soares

**Supplementary Tables and Figures**

### Supplementary Tables

### Supplementary Figures

Supplementary Table 1. Description of EPDS timepoints for women included in the main analysis.

| ALSPAC Questionnaire Name | Median Year of Completion | Mean age (years) | Median EPDS (IQR) | N |
| --- | --- | --- | --- | --- |
| G | 1993 | 33.7 (SD = 4, range: 20-45) | 4 (2 - 8) | 1959 |
| H | 1995 | 35.8 (SD = 4, range: 22-47) | 5 (2 - 9) | 1925 |
| K | 1997 | 37.5 (SD = 4, range: 24-49) | 5 (2 - 9) | 1901 |
| L | 1998 | 38.6 (SD = 3.9, range: 25-49) | 5 (2 - 9) | 1876 |
| N | 2000 | 41.2 (SD = 3.9, range: 28-52) | 5 (2 - 9) | 1820 |
| R | 2003 | 43.7 (SD = 3.9, range: 30-56) | 5 (2 - 9) | 1856 |
| T | 2010 | 51.6 (SD = 3.9, range: 37-63) | 6 (3 - 11) | 1439 |
| V | 2013 | 54.2 (SD = 3.9, range: 40-66) | 5 (2 - 10) | 1582 |
| Y | 2020 | 60.5 (SD = 4, range: 47-72) | 7 (3 - 11) | 1523 |
| Covid V | 2021 | 62 (SD = 4, range: 48-74) | 6 (2 - 11) | 1515 |
| MB | 2023 | 63.4 (SD = 4, range: 50-75) | 5 (2 - 10) | 1512 |

ALSPAC: Avon Longitudinal Study of Parents and Children

EPDS: Edinburgh Postnatal Depression Scale

SD: Standard Deviation

Supplementary Table 2. Derivation of the EPDS depressive symptom score outcome variable from Avon Longitudinal Study of Parents and Children (ALSPAC).

| **Data collection** | **ALSPAC Variable** | **Question(s)/measure(s)** | **Responses** | **Final variable coding** |
| --- | --- | --- | --- | --- |
| **EPDS** | | | | |
|  |  | **In the past week** | | Recode  (a), (b), (d): (1 = 0) (2 = 1) (3 = 2) (4 = 3)  (c),(e),(f),(g),(h),(i),(j): (1 = 3) (2 = 2) (3 = 1) (4 = 0)  Summed scored then calculated  **Mode imputation:** Any missing items were put to the mode for that item. If all items were missing, score was set as NA.  (Higher score = higher depressive symptom burden) |
| **Edinburgh Post Natal Depression Scale (EPDS) developed by Cox et al (1987).** | g280, h190, k3030, l2010, n6060, r4010, t3240, V5158, Y4000, covid5m_4000, MB6010, | (a) I have been able to laugh and see the funny side of things | 1: As much as I always could  2: Not quite so much now  3: Definitely not so much now  4:Not at all |  |
|  | g281, h191, k3031, l2011, n6061, r4011, t3241, V5159, Y4010, covid5m_4001, MB6020 | (b) I have looked forward with enjoyment to things | 1: As much as I ever did  2: Rather less than I used to  3: Definitely less than I used to  4: Hardly at all |  |
|  | g282, h192, k3032, l2012, n6062, r4012, t3242, V5150, Y4020, covid5m_4002, MB6030 | (c) I have blamed myself unnecessarily when things went wrong | 1: Yes, most of the time  2: Yes, some of the time  3: Not very often  4: No never |  |
|  | g283, h193, k3033, l2013, n6063, r4013, t3243, V5151, Y4030, covid5m_4003, MB6040 | (d) I have been anxious or worried for no good reason | 1: No, not at all  2: Hardly ever  3: Yes, sometimes  4: Yes, often |  |
|  | g284, h194, k3034, l2014, n6064, r4014, t3244, V5152, Y4040, covid5m_4004, MB6050 | (e) I have felt scared or panicky for no very good reason | 1: Yes, quite a lot  2: Yes, sometimes  3: No, not much  4: No, not at all |  |
|  | g285, h195, k3035, l2015, n6065, r4015, t3245, V5153, Y4050, covid5m_4005, MB6060 | (f) Things have been getting on top of me | 1: Yes, most of the time I haven't been able to cope  2: Yes, sometimes I haven't been coping as well as usual  3: No, most of the time I have coped quite well  4: No, I have been coping as well as ever |  |
|  | g286, h196, k3036, l2016, n6066, r4016, t3246, V5154, Y4060, covid5m_4006, MB6070 | (g) I have been so unhappy that I have had difficulty sleeping | 1: Yes, most of the time  2: Yes, sometimes  3: Not very often  4: No, not at all |  |
|  | g287, h197, k3037, l2017, n6067, r4017, t3247, V5155, Y4070, covid5m_4007, MB6080 | (h) I have felt sad or miserable | 1: Yes, most of the time  2: Yes, quite often  3: Not very often  4: No, not at all |  |
|  | g288, h198, k3038, l2018, n6068, r4018, t3248, V5156, Y4080, covid5m_4008, MB6090 | (i) I have been so unhappy that I have been crying | 1: Yes, most of the time  2: Yes, quite often  3: Only occasionally  4: No, never |  |
|  | g289, h199, k3039, l2019, n6069, r4019, t3249, V5157, Y4090, covid5m_4009, MB6100 | (j) The thought of harming myself has occurred to me | 1: Yes, quite often  2: Sometimes  3: Hardly ever  4: Never |  |

Supplementary Table 3. Derivation of the confounder variables from Avon Longitudinal Study of Parents and Children (ALSPAC).

| **Data collection** | **ALSPAC Variable** | **Question(s)/measure(s)** | **Responses** | **Final variable coding** |
| --- | --- | --- | --- | --- |
| **Age** | | | | |
|  | g994, h992, k9996a, l9996a, n9992, r9996a, t9994, V9996, Y9992, covid5m_9650, MB9510 | Derived from questionnaire date of completion and participants date of birth | Day/Month/Year | Age (years) |
| **Social Class (antenatal questionnaire)** | | | | |
| 1991 British Office of Population and Census Statistics job codes | C755 | Derived variable from questions:   - Actual job, occupation, trade or profession - Please tick which of the following apply to you: foreman, manager, supervisor, leading hand, self-employed, none of these   Type of industry or service given (main things done in job) | 1: I  2: II  3: III (non-manual)  4: IV (manual)  5: V  6: VI  65: Armed forces  -1: missing | Select highest social class between participant and their partner. Armed forces coded as missing. Coded from 1 (highest) to 6 (lowest).  Definitions:  Non-manual: professional, managerial, or skilled professions  Manual: partly or unskilled occupations |
|  | C765 |  |  |  |
| **Ethnicity (antenatal questionnaire)** | | | | |
|  | c800, n4180 | How would you describe the race or ethnic group of yourself? | -1 Missing  1 White  2 Black Caribbean  3 Black African  4 Other black  5 Indian  6 Pakistani  7 Bangladeshi  8 Chinese  9 Other  99 DK | Recode:  0: White  1: Non-white  Missing values from Questionnaire C were replaced using responses from Questionnaire N |
| **Education** | | | | |
|  | c645a | Derived variable: mother’s highest educational qualification | 1: CSE/none  2: Vocational  3: O-level  4: A-level  5: Degree  -1: Missing | Recode:  0: A-level or greater  1: O-level  2: Vocational or less  Education variables were not derived by ALSPAC for Questionnaires K and N. The same coding as applied in questionnaire C was used.  The highest educational qualification reported was used. It was felt that a lot of mothers with no educational qualifications left this whole question blank. Therefore, all women who did not respond to any of the questions were coded to 2 (Vocational or less) |
|  | k6280, k6281 | CSE/none |  |  |
|  | k6284, k6285, k6288, k6295 | Vocational |  |  |
|  | k6282, | O-level |  |  |
|  | k6283, k6286, k6287, k6289, k6290, k6291 | A-level |  |  |
|  | k6292 | Degree |  |  |
|  | n4012, n4000 | CSE/none |  |  |
|  | n4003, n4004, n4007, n4015 | Vocational |  |  |
|  | n4001 | O-level |  |  |
|  | n4002, n4005, n4006, n4008, n4009, n4010 | A-level |  |  |
|  | n4011 | Degree |  |  |
| **Age at menarche** | | | | |
|  | d010a, n1120, r2080 | How old were you when your periods first started | Age (years) | Recode:  0: 8 – 11 years  1: 12 – 14 years  2: 15 or older  Missing responses from Questionnaire D were replaced from Questionnaire N and Questionnaire R |
| **Material hardship** | | | | |
| ALSPAC hardship items |  | **How difficult at the moment do you find it to afford these items:** | |  |
|  | c520 , f800 ,g835, h730, k6200, m5170 ,t1360 | a) food | 1: Very difficult  2: Fairly difficult  3: Slightly difficult  4: Not difficult  5: Paid directly by social security | Recode responses of 5 (Paid directly by social security) to 4  Material hardship = 20 – summed scored of 5 questions.  (Higher scores = more material hardship) |
|  | c521, f801, g836, h731, k6201, m5171, t1361 | b) clothing |  |  |
|  | c522, f802, g837, h732, k6202, m5172, t1362 | c) heating |  |  |
|  | c523, f803, g838, h733, k6203, m5173, t1363 | d) rent or mortgage |  |  |
|  | c524, f804, g839, h734, k6204, m5174, t1364 | e) things you need for your children |  |  |
| **Social support** | | | | |
| ALSPAC social support items | d790, e600, f910, g216, k8020, l7020, p4020, s6020 | a) Mother feels she has no-one to share feelings with | 1: Exactly feel  2: Often feel  3: Sometimes feel  4: Never feel | (a), (e): (1=0)(2=1)(3=2)(4=3)  (b), (c), (d), (f), (g), (h), (i), (j): (1=3)(2=2)(3=1)(4=0)  Variable scores then summed for a maximum of 30 (higher score = more social support)  Mode imputation: missing values were put to the mode for that variable. |
|  | d791, e601, f911, g217, k8021, l7021, p4021, s6021 | b) Mother feels her partner provides the emotional support she needs |  |  |
|  | d792, e602, f912, g218, k8022, l7022, p4022, s6022 | c) Mother can share experiences with other mothers |  |  |
|  | d793, e603, f913, g219, k8023, l7023, p4023, s6023 | d) Mother feels her neighbours would help in moments of difficulty |  |  |
|  | d794, e604, f914, g220, k8024, l7024, p4024, s6024 | e) Mother is worried that partner might leave |  |  |
|  | d795, e605, f915, g221, k8025, l7025, p4025, s6025 | f) Mother always has someone to share happiness about child |  |  |
|  | d796, e606, f916, g222, k8026, l7026, p4026, s6026 | g) Partner will take over from mother if she is tired |  |  |
|  | d797, e607, f917, g223, k8027, l7027, p4027, s6027 | h) Mothers family would help in financial difficulty |  |  |
|  | d798, e608, f918, g224, k8028, l7028, p4028, s6028 | i) Mothers friends would help in financial difficulty |  |  |
|  | d799, e609, f919, g225, k8029, l7029, p4029, s6029 | j) Mother feels if all fails state would support her financially |  |  |
| **Smoking status** | | | | |
|  | b650, n5000, n5002, n5003, n5004, n5005, r6010, r6012, r6013, r6014, r6015, t5560, MA8010, MA8070 | Ever smoked | Yes/No | Recoded into Never, Ever & Current smoker.  Never and Ever smoker responses were sense checked with historical responses. |
|  | f621, g820, h720, j735, k6180, l5050, l5051, m5160, n5010, r6020, s1300, s1301, t5526, V5526, MA8030, MA8060 | No smoked daily at PRES | None, or daily cigarette brackets (e.g. 1–4 to 30+) |  |
|  | s1302, s1303, t5520, t5521, V5520, V5521, MA8080, n5008, r6018, MA8040 | Mother currently smokes | Yes/No |  |
| **Body mass index (BMI)** | | | | |
|  | dw021, m4221, n1145, p1291, s1291, V4400, XB070, MB4540, fm1ms100, fm2ms100, fm3ms100, fm4ms100 | Height | Metres | Average height across all responses was calculated. BMI (kg/m^2^) |
|  | dw002, m4220, n1140, p1290, s1290, V4410, XB080, MB4580, fm1ms110, fm2ms110, fm3ms110, fm4ms110 | Weight | kg |  |
| **Alcohol intake** | | | | |
|  | f626, g825, h724, k6191, t5500, V5500, MA8220 | How many days in the past month did you have the equivalent of 2 pints of beer, 4 glasses of wine or 4 pub measures of spirit? | 1: Every day  2: more than 10 days  3: 5-10 days  4: 3-4 days  5: 1-2 days  6: none | Recode (4/6 = 0) (3=1) (1/2 =2)  0: Never or ≤4 times/month  1: 2–3 times/week  2: ≥ 4 times/week |
| **Hormone replacement therapy (HRT)** | | | | |
|  | p1055, q4293, q4295, s4293, s4295 | Frequency mother has taken hormone replacement therapy in last 2 years | 1: Every day  2: Often  3: Sometimes  4: Not at all  5: Once | Recoded to Never vs Ever HRT users |
|  | q4290, s4290, t4920 | Mother used medicine in last 12 months for HRT | Yes/No |  |
|  | t4961, V4955, Y5140, MB4790, MB4800, MB4810, MB4820, fm1sa206, fm1ob110, fm2ob110a, fm2ob110b, fm2ob110c, fm3ob110a, fm3ob110b, fm3ob110c, fm4ob110a, fm4ob110b, fm4ob110c | Currently on HRT | Yes/No |  |
|  | t4960, V4954 | Respondent has stopped HRT and then started again | Yes/No |  |
|  | V4920, Y5130 | Respondent ever had hormone replacement therapy (HRT) | Yes/No |  |
| **History of Baseline Depression** | | | | |
|  | d171 | Have you ever had any of the following problems:  Severe depression | 1: Yes had it recently  2: Yes in past, not now  3: No never | Recode (1/2 = 1) (3=0)  0: No history of baseline depression  1: History of baseline depression |

Supplementary Table 4. Comparison of model fit across linear, quadratic and restricted cubic spline models.

|  | Corresponding Figure | N participants | N observations | df | AIC | BIC |
| --- | --- | --- | --- | --- | --- | --- |
| Main analysis |  |  |  |  |  |  |
| Trajectories of depressive symptoms in the years surrounding the FMP | Figure 2a | 2,036 | 18,466 | Linear | 103656.1 | 103867.3 |
|  |  |  |  | Quadratic | 103657.7 | 103876.8 |
|  |  |  |  | 2 | 103658.1 | 103877.1 |
|  |  |  |  | 3 | 103635.8 | 103862.6 |
|  |  |  |  | 4 | 103631.4 | 103866.1 |
|  |  |  |  | 5 | 103623.1 | 103865.7 |
| Trajectories of depressive symptoms across chronological age | Figure 2b | 2,036 | 18,466 | Linear | 103720.1 | 103915.7 |
|  |  |  |  | Quadratic | 103658.7 | 103862.1 |
|  |  |  |  | 2 | 103668.9 | 103872.3 |
|  |  |  |  | 3 | 103639.7 | 103850.9 |
|  |  |  |  | 4 | 103623.1 | 103842.2 |
|  |  |  |  | 5 | 103623.0 | 103849.9 |
| Trajectories of depressive symptoms across chronological age by age at menopause categories | Figure 4 | 2,036 | 18,466 | Linear | 103723.7 | 103935.0 |
|  |  |  |  | Quadratic | 103662.4 | 103889.3 |
|  |  |  |  | 2 | 103673.0 | 103899.8 |
|  |  |  |  | 3 | 103645.2 | 103887.8 |
|  |  |  |  | 4 | 103630.1 | 103888.3 |
|  |  |  |  | 5 | 103631.2 | 103905.1 |

Supplementary Table 5. Overview of Multilevel Model Structures and Covariate Adjustments. All models included a participant level random intercept.

| **Main exposure** | **Exposure Modelling Approach** | **Outcome** | **Fixed effect (Confounders)** | **Random Effects** |
| --- | --- | --- | --- | --- |
| Figure 2a | | | | |
| Reproductive age | Linear | EPDS (continuous); modelled with Normal distribution | Centred age, centred age squared, ethnicity, social class, education, age at menarche, material hardship, social support, smoking status, BMI, alcohol intake | Random slope for chronological age |
| Figure 2b | | | | |
| Chronological age | Quadratic (age and age^2^) | EPDS (continuous); modelled with Normal distribution | Ethnicity, social class, education, age at menarche, material hardship, social support, smoking status, BMI, alcohol intake | Random slope for chronological age |
| Figure 3 | | | | |
| Chronological age | Quadratic (age and age²); interaction with age at menopause category | EPDS (continuous); modelled with Normal distribution | Ethnicity, social class, education, age at menarche, material hardship, social support, smoking status, BMI, alcohol intake | Random slope for chronological age |
| Figure 4 | | | | |
| Chronological age | Linear; interaction with menopausal stage | EPDS (continuous); modelled with Normal distribution | Ethnicity, social class, education, age at menarche, material hardship, social support, smoking status, BMI, alcohol intake | Random slope for chronological age |
| Figure 5a | | | | |
| Reproductive age | Linear | EPDS (binary); modelled with Binomial distribution | Centred age, centred age squared, ethnicity, social class, education, age at menarche, material hardship, social support, smoking status, BMI, alcohol intake | Random slope for chronological age |
| Figure 5b | | | | |
| Chronological age | Quadratic (age and age^2^) | EPDS (binary); modelled with Binomial distribution | Ethnicity, social class, education, age at menarche, material hardship, social support, smoking status, BMI, alcohol intake | Random slope for chronological age |
| Supplementary Figure 1 | | | | |
| Reproductive age | (a) Linear | EPDS (continuous); modelled with Normal distribution | Centred age, centred age squared, ethnicity, social class, education, age at menarche, material hardship, social support, smoking status, BMI, alcohol intake | Random slope for chronological age |
|  | (b) Quadratic (age and age²) |  |  |  |
|  | (c) Restricted cubic spline (3 knots) |  |  |  |
|  | (d) Restricted cubic spline (4 knots) |  |  |  |
|  | (e) Restricted cubic spline (5 knots) |  |  |  |
|  | (f) Restricted cubic spline (6 knots) |  |  |  |
| Supplementary Figure 2 | | | | |
| Chronological age | (a) Linear | EPDS (continuous); modelled with Normal distribution | Ethnicity, social class, education, age at menarche, material hardship, social support, smoking status, BMI, alcohol intake | Random slope for chronological age |
|  | (b) Quadratic (age and age²) |  |  |  |
|  | (c) Restricted cubic spline (3 knots) |  |  |  |
|  | (d) Restricted cubic spline (4 knots) |  |  |  |
|  | (e) Restricted cubic spline (5 knots) |  |  |  |
|  | (f) Restricted cubic spline (6 knots) |  |  |  |
| Supplementary Figure 3 | | | | |
| Reproductive age | Linear | EPDS (continuous); modelled with Normal distribution | Ethnicity, social class, education, age at menarche, material hardship, social support, smoking status, BMI, alcohol intake | Random slope for reproductive age |
| Supplementary Figure 4 | | | | |
| Chronological age; interaction for menopausal stages | Linear; interaction with menopausal stages | EPDS (binary); modelled with Binomial distribution | Ethnicity, social class, education, age at menarche, material hardship, social support, smoking status, BMI, alcohol intake | Random slope chronological age |
| Supplementary Figure 5 | | | | |
| Chronological age; interaction for age at menopause categories | Quadratic (age and age^2^); interaction with menopausal age | EPDS (binary); modelled with Binomial distribution | Ethnicity, social class, education, age at menarche, material hardship, social support, smoking status, BMI, alcohol intake | Random slope chronological age |
| Supplementary Figure 6a | | | | |
| Reproductive age | Linear | EPDS (continuous); modelled with Normal distribution | Centred age, centred age^2^ , ethnicity, social class, education, age at menarche, material hardship, social support, smoking status, BMI, alcohol intake | Random slope for chronological age |
| Supplementary Figure 6b | | | | |
| Chronological age | Quadratic (age and age^2^) | EPDS (continuous); modelled with Normal distribution | Ethnicity, social class, education, age at menarche, material hardship, social support, smoking status, BMI, alcohol intake | Random slope for chronological age |
| Supplementary Figure 7a | | | | |
| Reproductive age | Linear | EPDS (continuous – log transformed); modelled with Normal distribution | Centred age, centred age^2^ , ethnicity, social class, education, age at menarche, material hardship, social support, smoking status, BMI, alcohol intake | Random slope for chronological age |
| Supplementary Figure 7b | | | | |
| Chronological age | Quadratic (age and age^2^) | EPDS (continuous - log transformed); modelled with Normal distribution | Ethnicity, social class, education, age at menarche, material hardship, social support, smoking status, BMI, alcohol intake | Random slope for chronological age |
| Supplementary Figure 8a | | | | |
| Reproductive age | Linear | EPDS (continuous); modelled with Normal distribution | Centred age, centred age^2^ , ethnicity, social class, education, age at menarche, material hardship, social support, smoking status, BMI, alcohol intake, history of depression | Random slope for chronological age |
| Supplementary Figure 8b | | | | |
| Chronological age | Quadratic (age and age^2^) | EPDS (continuous); modelled with Normal distribution | Ethnicity, social class, education, age at menarche, material hardship, social support, smoking status, BMI, alcohol intake, history of depression | Random slope for chronological age |
| Supplementary Figure 9a | | | | |
| Reproductive age | Linear | EPDS (binary); modelled with Binomial distribution | Centred age, centred age^2^ , ethnicity, social class, education, age at menarche, material hardship, social support, smoking status, BMI, alcohol intake, history of depression | Random slope for chronological age |
| Supplementary Figure 9b | | | | |
| Chronological age | Quadratic (age and age^2^) | EPDS (binary); modelled with Binomial distribution | Ethnicity, social class, education, age at menarche, material hardship, social support, smoking status, BMI, alcohol intake, history of depression | Random slope for chronological age |
| Supplementary Figure 10 | | | | |
| Chronological age | Quadratic (age and age^2^) | EPDS (continuous); modelled with Normal distribution | Ethnicity, social class, education, age at menarche, material hardship, social support, smoking status, BMI, alcohol intake | Random slope for chronological age |
| Supplementary Figure 11 | | | | |
| Reproductive age; interaction for HRT use | Linear; interaction for HRT use | EPDS (continuous); modelled with Normal distribution | Centred age, centred age^2^ , ethnicity, social class, education, age at menarche, material hardship, social support, smoking status, BMI, alcohol intake | Random slope for chronological age |

Supplementary Table 6. Average predicted depressive symptom scores across reproductive age.

| Years since the final menstrual period | Predicted depressive symptom score (95% CI), adjusted for chronological age | Predicted depressive symptom score (95% CI), unadjusted for chronological age |
| --- | --- | --- |
| -20 | 5.90 (4.88, 6.92) | 5.51 (5.08, 5.94) |
| -15 | 6.15 (5.33, 6.97) | 5.75 (5.32, 6.17) |
| -10 | 6.40 (5.75, 7.05) | 5.98 (5.55, 6.41) |
| -5 | 6.65 (6.14, 7.16) | 6.22 (5.78, 6.66) |
| 0 | 6.90 (6.44, 7.36) | 6.46 (6.01, 6.90) |
| 5 | 7.15 (6.63, 7.66) | 6.69 (6.23, 7.16) |
| 10 | 7.40 (6.75, 8.05) | 6.93 (6.45, 7.42) |
| 15 | 7.65 (6.82, 8.47) | 7.17 (6.66, 7.68) |
| 20 | 7.90 (6.87, 8.92) | 7.41 (6.87, 7.94) |

Predictions are made for a woman aged 50 years. Model was adjusted for ethnicity, social class, age at menarche, education, material hardship, social support, smoking status, BMI, and alcohol intake.

Supplementary Table 7. Average predicted depressive symptom scores across chronological age.

| Age (years) | Predicted depressive symptom score (95% CI) |
| --- | --- |
| 30 | 5.11 (4.67, 5.56) |
| 35 | 5.73 (5.3, 6.16) |
| 40 | 6.23 (5.79, 6.66) |
| 45 | 6.62 (6.17, 7.07) |
| 50 | 6.9 (6.44, 7.36) |
| 55 | 7.06 (6.59, 7.53) |
| 60 | 7.11 (6.63, 7.6) |
| 65 | 7.05 (6.54, 7.56) |
| 70 | 6.88 (6.33, 7.43) |

Model was adjusted for ethnicity, social class, age at menarche, educational attainment, material hardship, social support, smoking status, BMI, and alcohol intake.

Supplementary Table 8. Association between menopause stage and depression, further adjusting for baseline history of depression as a confounder.

| Menopausal stage | Odds ratio | 95% C.I. | p-value |
| --- | --- | --- | --- |
| Reproductive (ref) | 1.00 (ref) | - | - |
| Perimenopause | 1.20 | 1.03 – 1.40 | 0.02 |
| Postmenopause | 1.00 | 0.83 – 1.21 | 0.98 |

Model adjusted for age, ethnicity, social class, education, age at menarche, material hardship, social support, smoking status, body mass index, alcohol intake and history of depression.

Supplementary Table 9. Comparison of confounders at baseline between women with and without an age at menopause.

| **Characteristic** | | **Age at menopause unavailable** | **Age at menopause available** | ***P*** |
| --- | --- | --- | --- | --- |
| N |  | 7,241 | 2,036 |  |
| Ethnicity (%) | White | 7,102 (98.1) | 1,991 (97.8) | 0.459 |
| Social class (%) | I (highest non-manual) | 209 ( 2.9) | 114 ( 5.6) | <0.001 |
|  | II (non-manual) | 1,533 (21.2) | 718 (35.3) |  |
|  | III (non-manual) | 1,839 (25.4) | 559 (27.5) |  |
|  | IV (manual) | 2,166 (29.9) | 413 (20.3) |  |
|  | V (manual) | 1,151 (15.9) | 190 ( 9.3) |  |
|  | VI (lowest manual) | 343 ( 4.7) | 42 ( 2.1) |  |
| Age at menarche (%) | Early (≤ 11 years) | 1,432 (19.8) | 340 (16.7) | 0.007 |
|  | Average (12-14 years) | 4,908 (67.8) | 1,426 (70.0) |  |
|  | Late (≥ 15 years) | 901 (12.4) | 270 (13.3) |  |
| Education (%) | CSE/vocational degree | 1,387 (19.2) | 216 (10.6) | <0.001 |
|  | O-level | 2,905 (40.1) | 579 (28.4) |  |
|  | A-level/University degree | 2,949 (40.7) | 1241 (61.0) |  |
| Smoking status (%) | Never | 3,657 (50.5) | 1,118 (54.9) | <0.001 |
|  | Ever | 1,917 (26.5) | 621 (30.5) |  |
|  | Current | 1,667 (23.0) | 297 (14.6) |  |
| Alcohol intake (%) | Never or less than 4 times a month | 6,246 (86.3) | 1,741 (85.5) | 0.69 |
|  | 2 to 3 times a week | 667 ( 9.2) | 198 ( 9.7) |  |
|  | 4 or more times a week | 328 ( 4.5) | 97 ( 4.8) |  |
| Material hardship (mean (SD)) | | 3.04 (3.53) | 2.29 (3.15) | <0.001 |
| Social support (mean (SD)) | | 20.44 (5.36) | 20.45 (5.27) | 0.938 |
| Body mass index (mean (SD)) | | 23.27 (4.08) | 22.72 (3.46) | <0.001 |

Supplementary Table 10. Proportion of observations with missing data for each confounder.

| **Confounder** | **Proportion of observations with missing value (%)** |
| --- | --- |
| Ethnicity | 2.1 |
| Social class | 8.63 |
| Education | 1.72 |
| Age at menarche | 4.64 |
| Material hardship | 1.87 |
| Social support | 0.32 |
| Smoking status | 0.01 |
| Body Mass Index | 2.76 |
| Alcohol intake | 0.91 |

Supplementary Figure 1. Comparison of model fit for trajectories of depressive symptoms in the years surrounding the final menstrual period. Reproductive age was modelled using: (a) a linear term, (b) quadratic term, and restricted cubic splines with (c) 2, (d) 3, (e) 4, and (f) 5 degrees of freedom. Dashed vertical line corresponds to menopause.

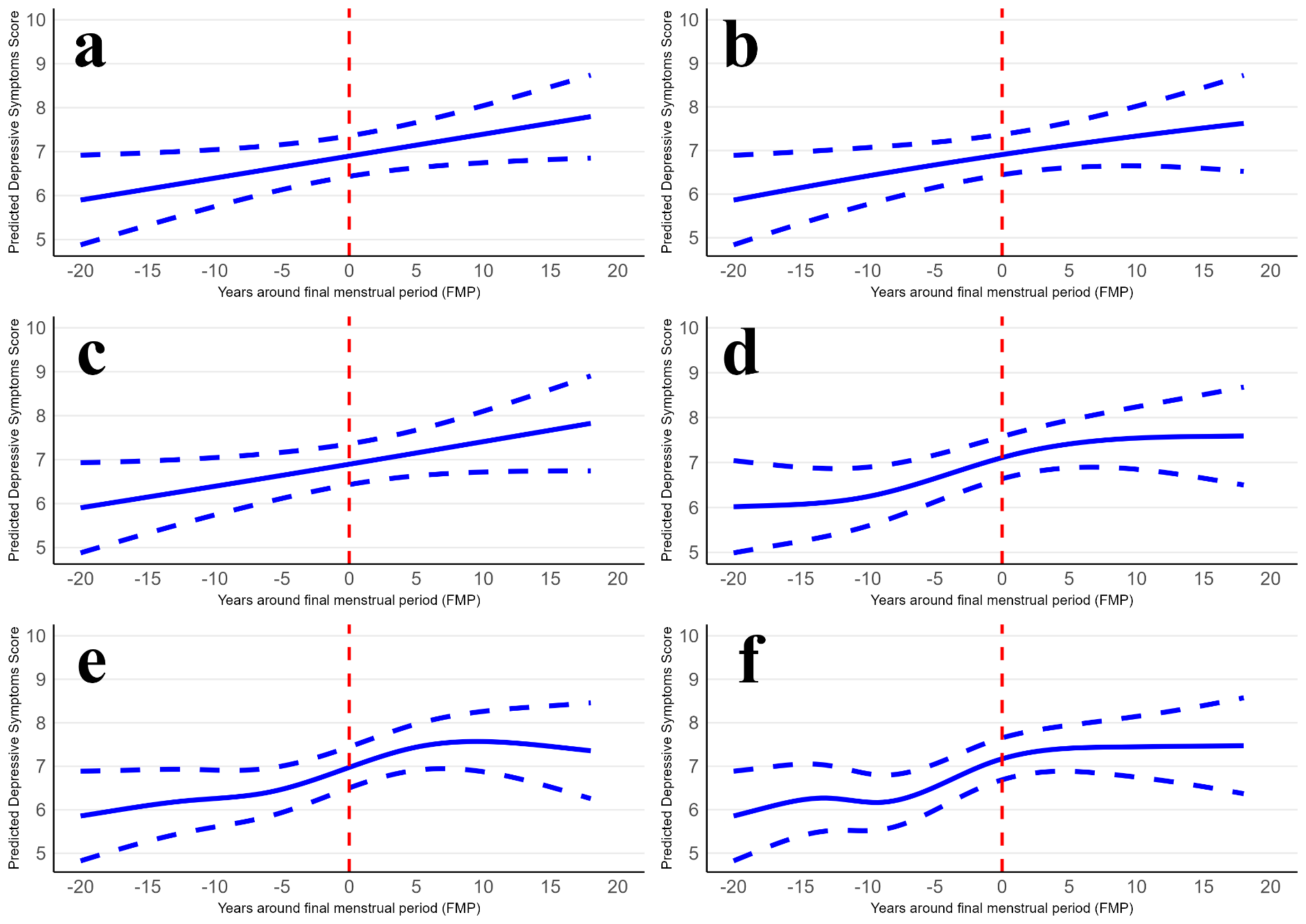

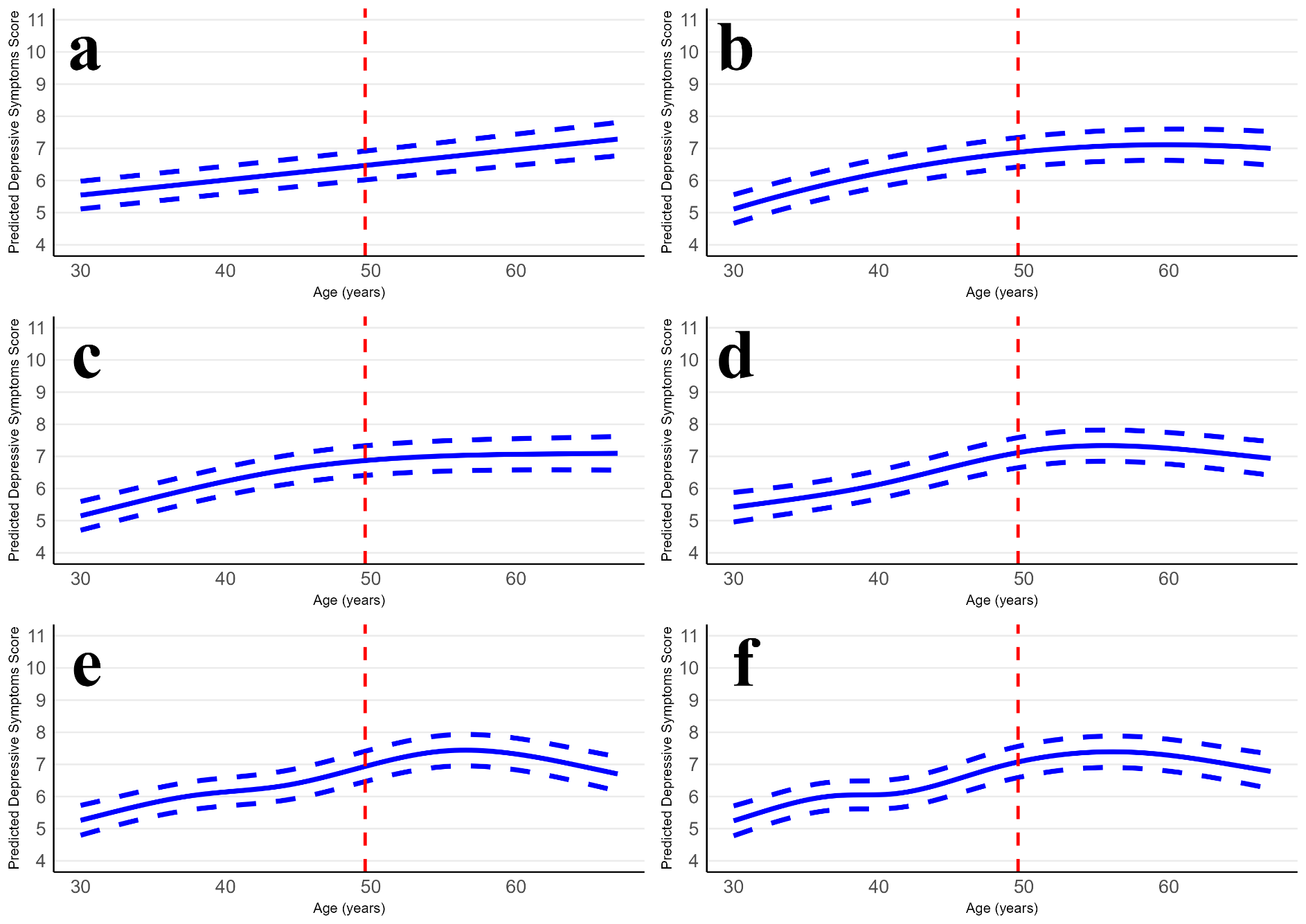
Supplementary Figure 2. Comparison of model fit for trajectories of depressive symptoms across chronological age. Chronological age was modelled using: (a) a linear term, (b) quadratic term, and restricted cubic splines with (c) 2, (d) 3, (e) 4, and (f) 5 degrees of freedom. Dashed vertical line corresponds to median age at menopause in the study.

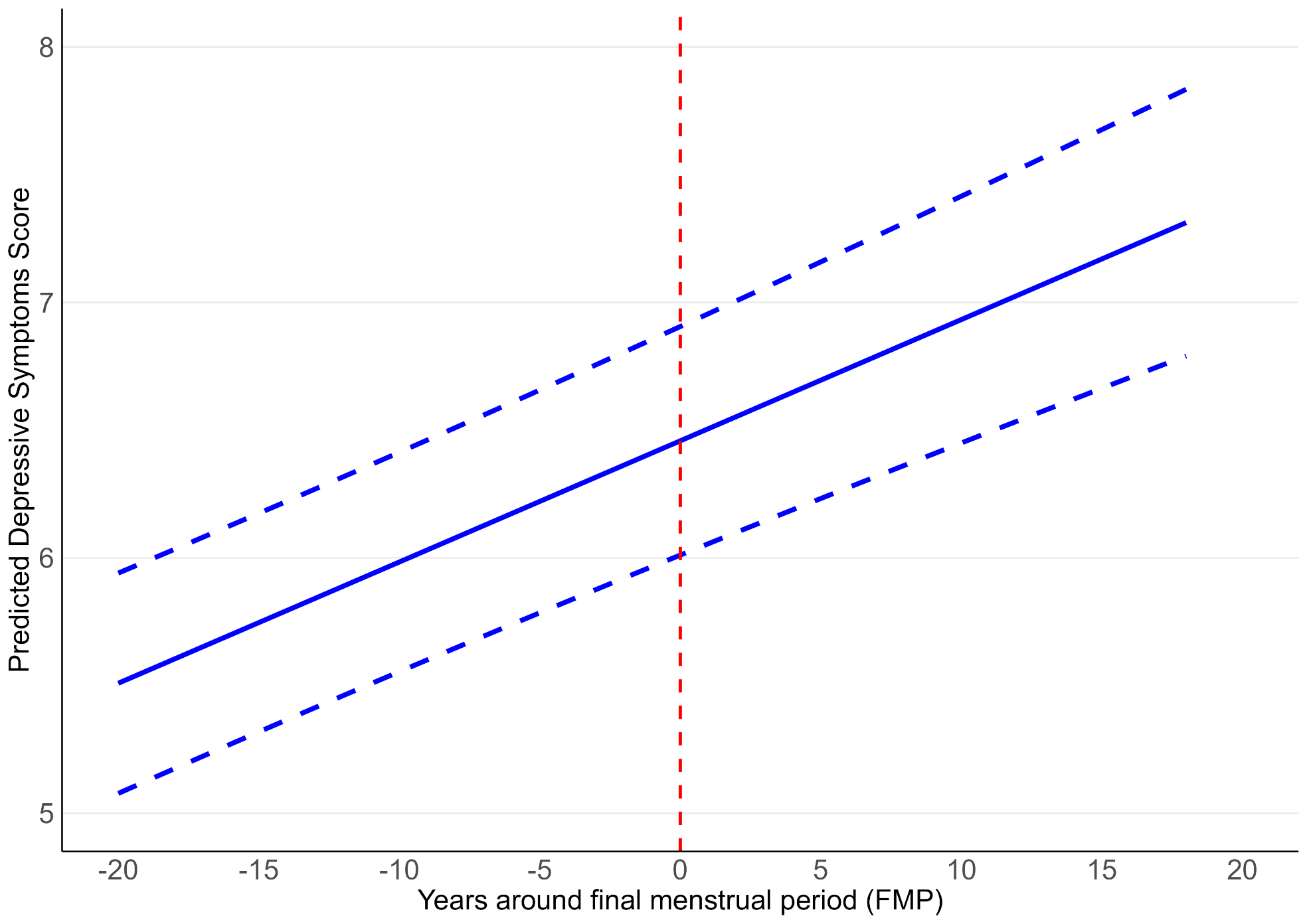
Supplementary Figure 3. Trajectory of depressive symptoms in the years surrounding the final menstrual period unadjusted for chronological age. Dashed vertical line corresponds to menopause.

Supplementary Figure 4. Predicted probability of depression across chronological age, with an interaction for menopausal stage (pre-, peri-, and post-menopause). Adjusted for ethnicity, social class, education, age at menarche, material hardship, social support, smoking status, BMI and alcohol intake.

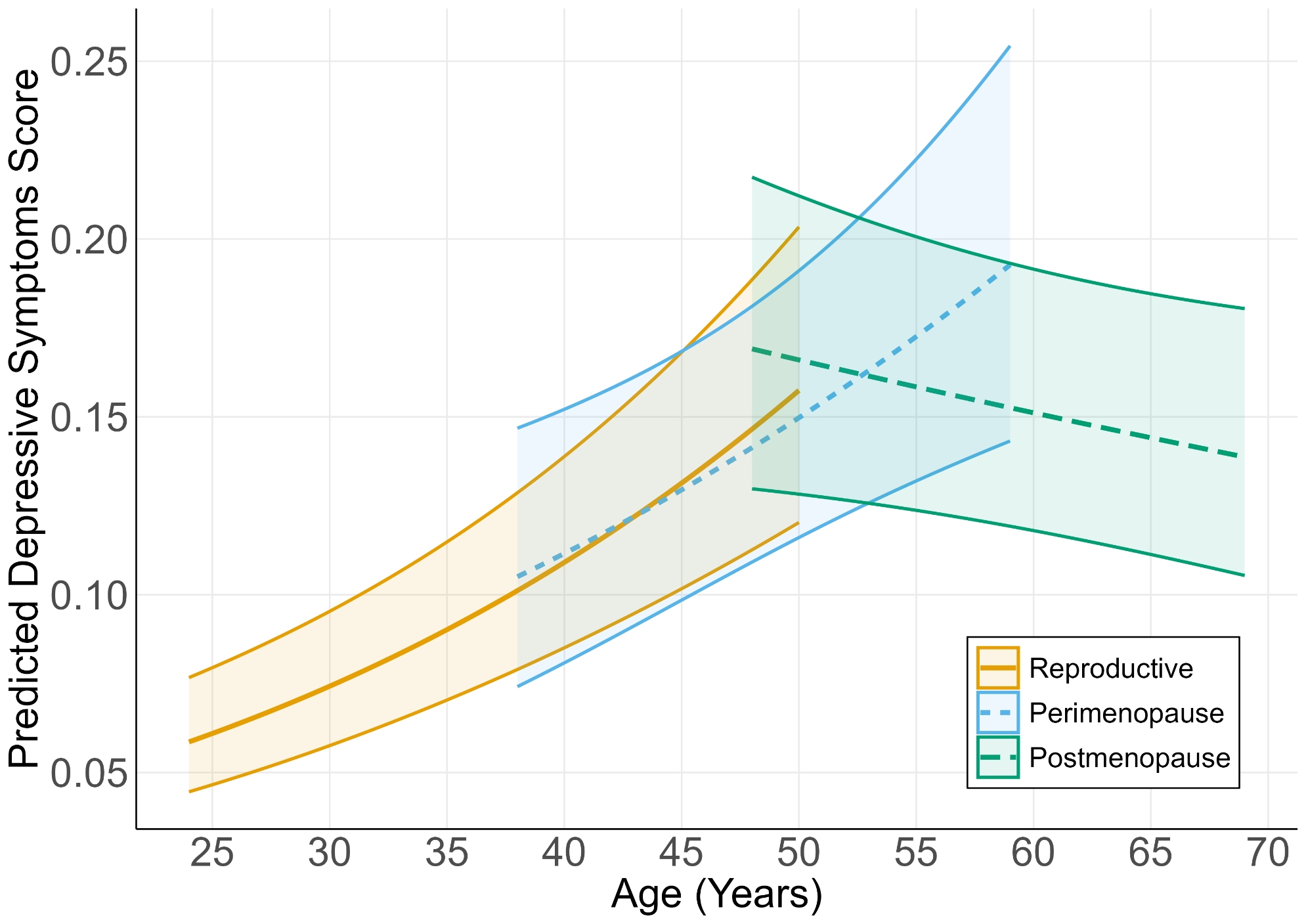

Supplementary Figure 5. Predicted probability of depression across chronological age, with an interaction for age at menopause categories. Adjusted for ethnicity, social class, education, age at menarche, material hardship, social support, smoking status, BMI and alcohol intake.

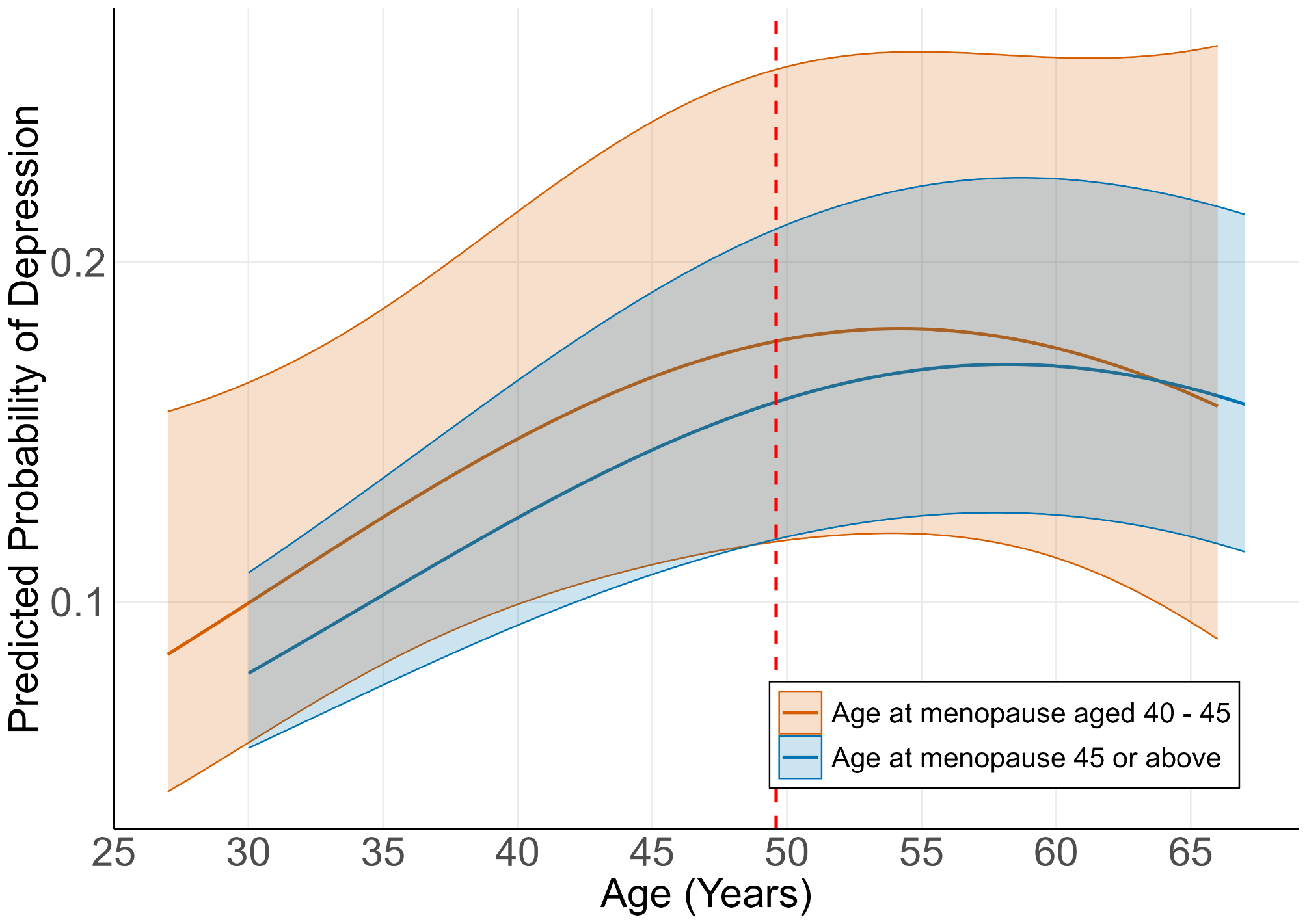

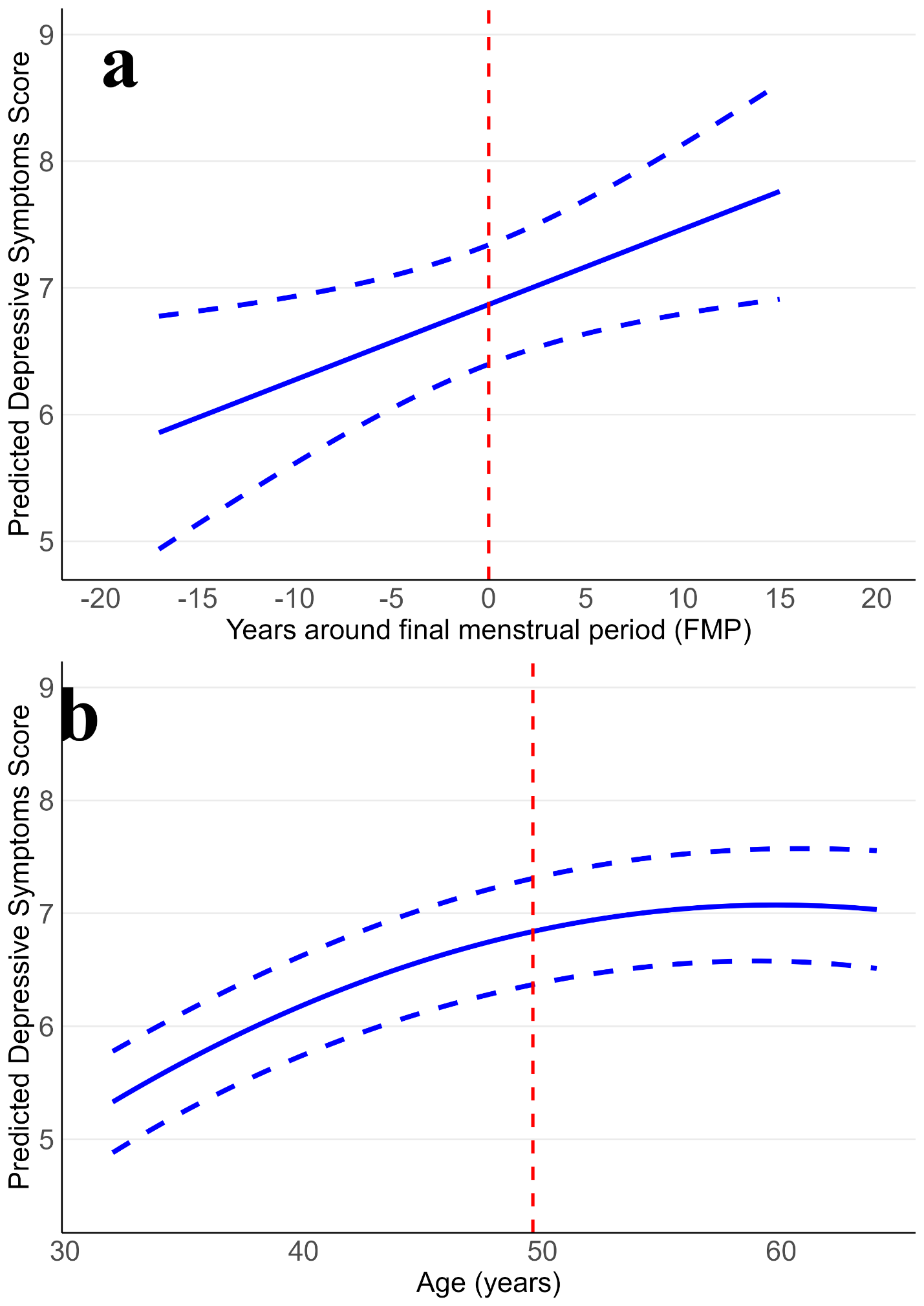
Supplementary Figure 6. Trajectories of depressive symptoms in the years surrounding the final menstrual period (A) and chronological age (B), restricted at the 5th and 95th centiles of age at final menstrual period. Dashed vertical line corresponds to menopause (A) or average age of menopause in the study (B).

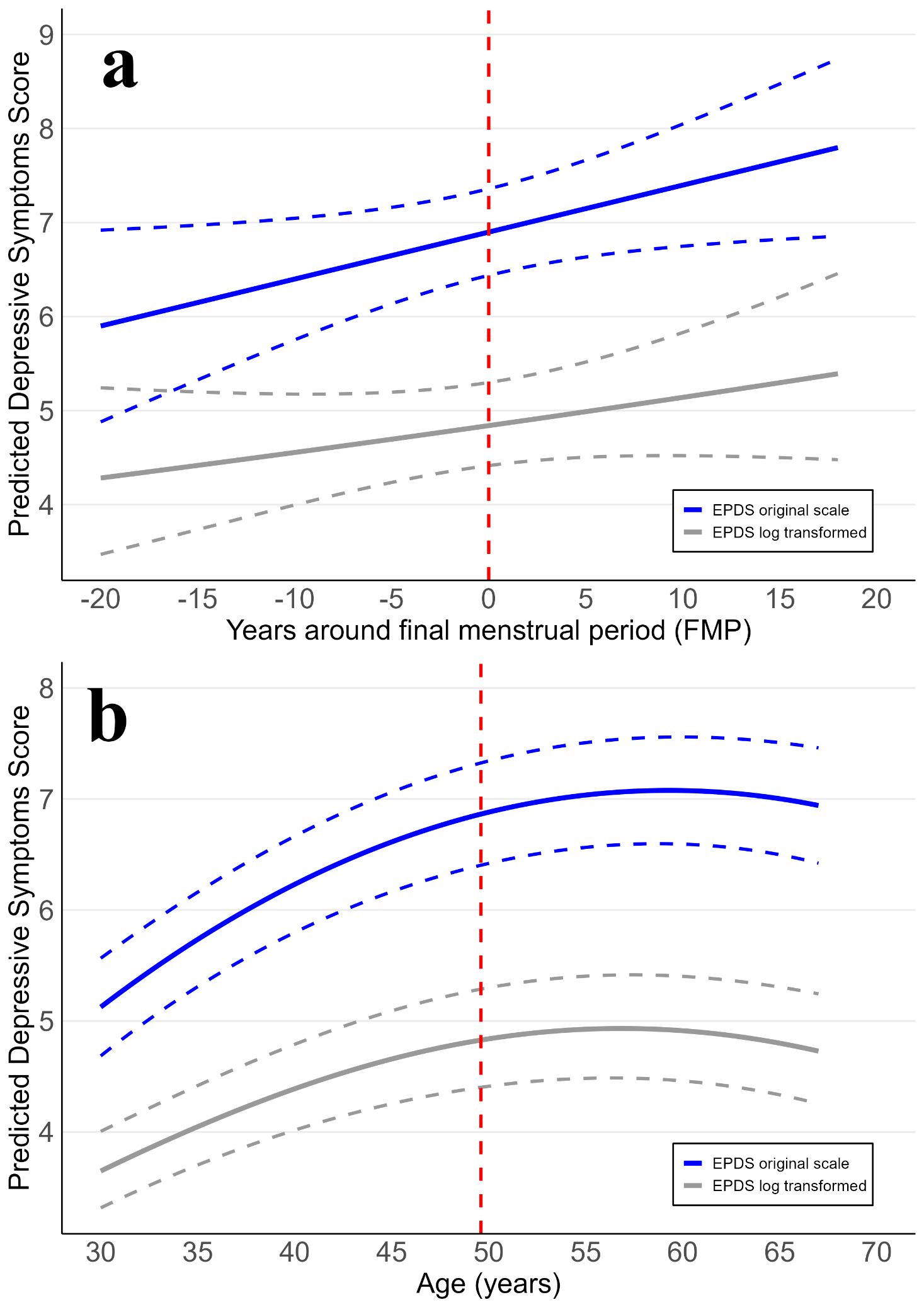
Supplementary Figure 7. Trajectories of depressive symptoms in the years surrounding the final menstrual period (A) and chronological age (B). EPDS score was log transformed prior to modelling. EPDS trajectories on the original scale are presented for comparison. Dashed vertical line corresponds to menopause (A) or average age of menopause in the study (B).

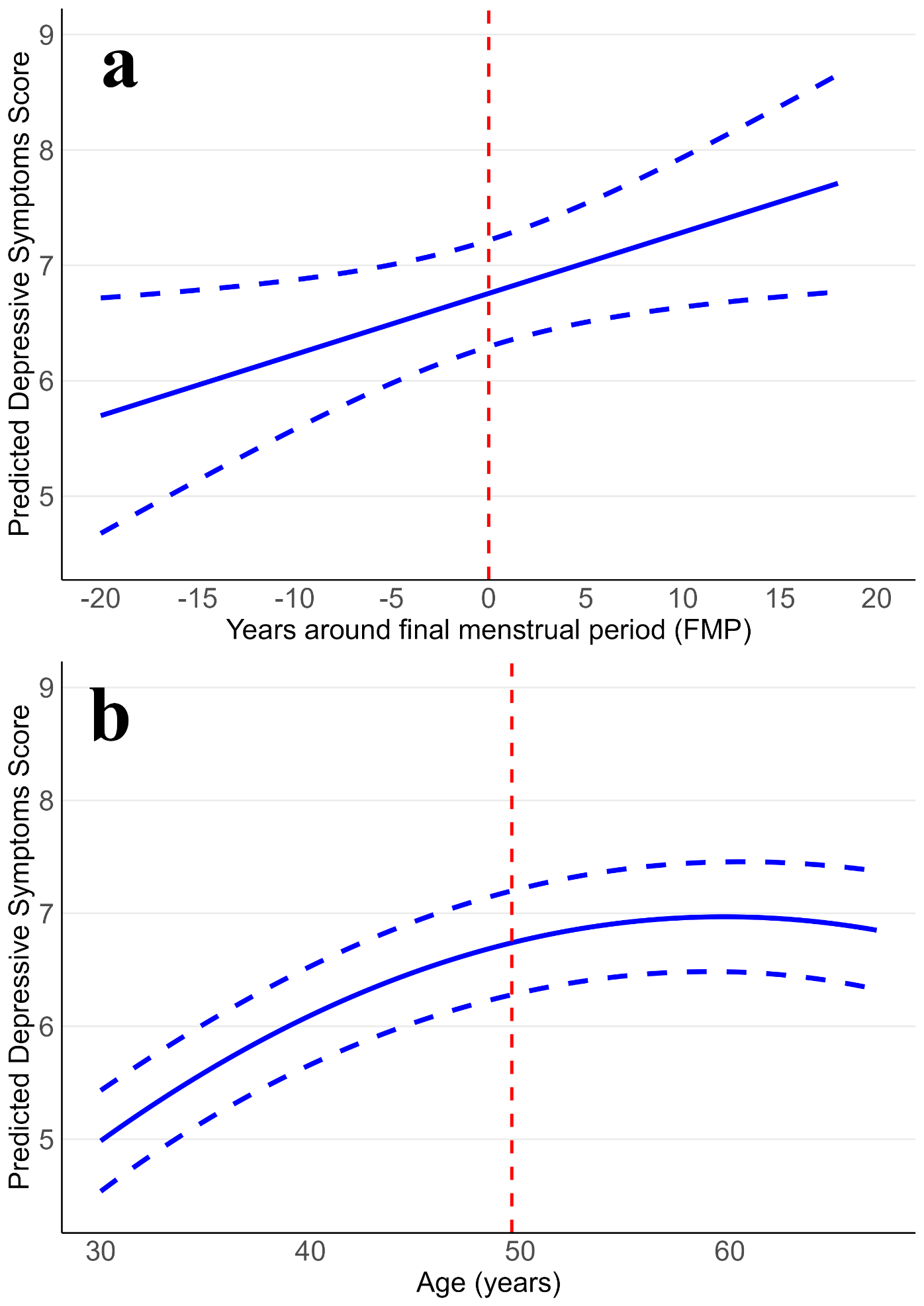
Supplementary Figure 8. Trajectories of depressive symptoms in the years surrounding the (A) final menstrual period and (B) chronological age, further adjusting for baseline history of depression as a confounder. Dashed vertical line corresponds to (A) menopause or (B) average age of menopause in the study.

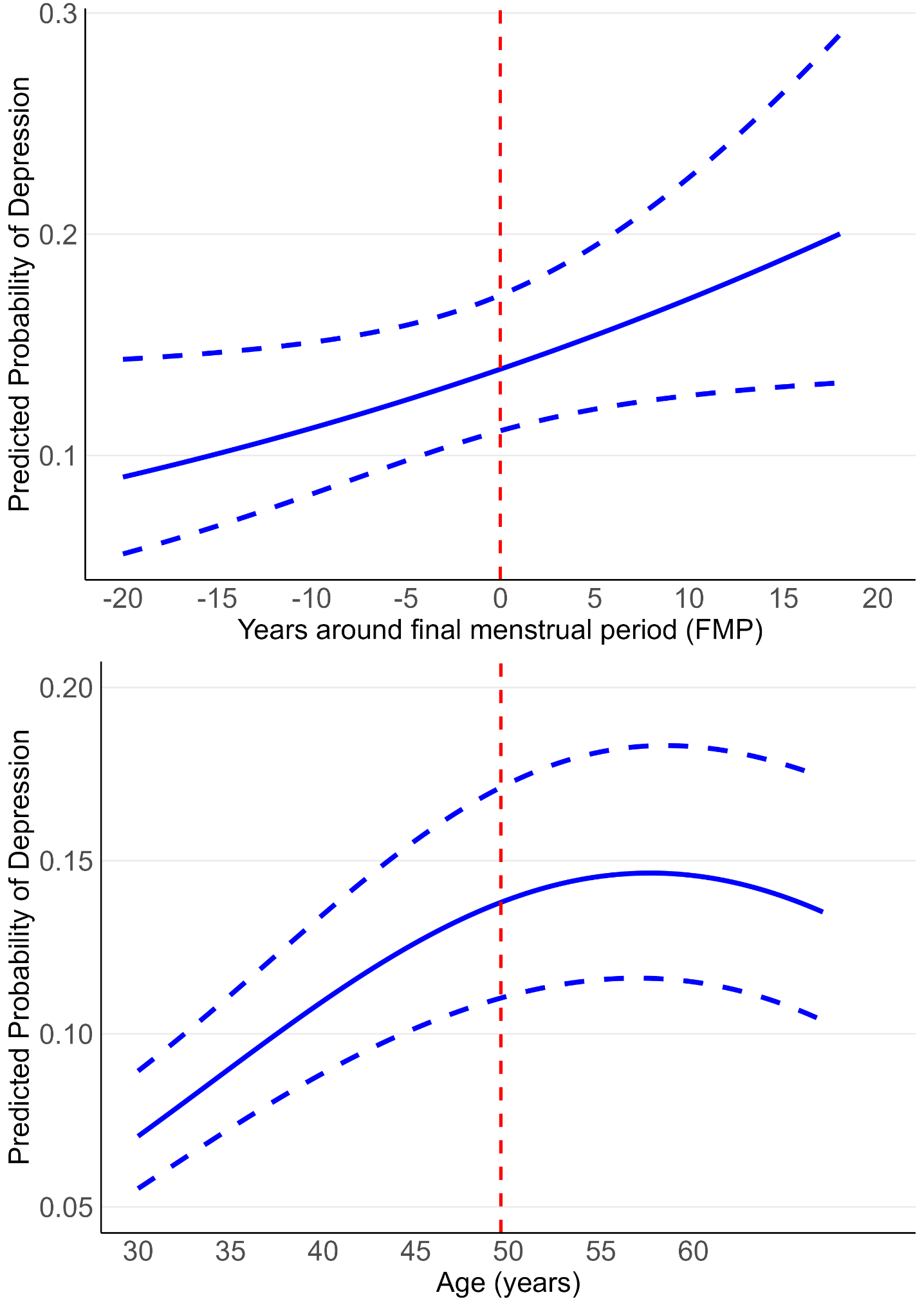
Supplementary Figure 9. Predicted probability of depression in the years surrounding the (A) final menstrual period and (B) chronological age, further adjusting for baseline history of depression as a confounder. Dashed vertical line corresponds to (A) menopause or (B) average age of menopause in the study.

Supplementary Figure 10. Comparison of trajectories of depressive symptoms across chronological age in the full population (N = 9,277) and the population with an age at menopause (N = 2,036). Model adjusted for all confounders. Dashed vertical line corresponds to average age at menopause in the study menopause.

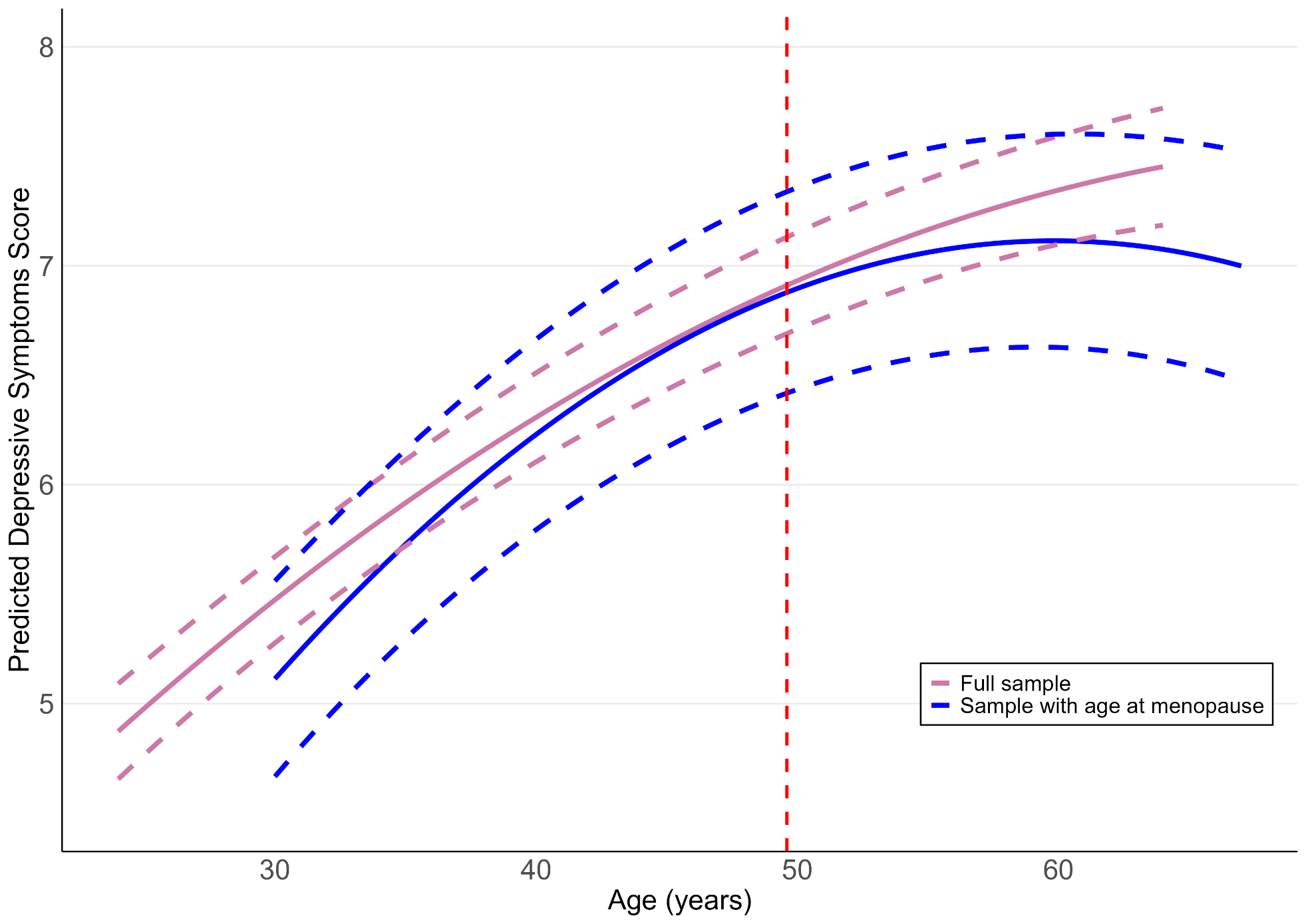

Supplementary Figure 11. Trajectories of depressive symptoms across reproductive age by HRT use (Ever vs. Never).

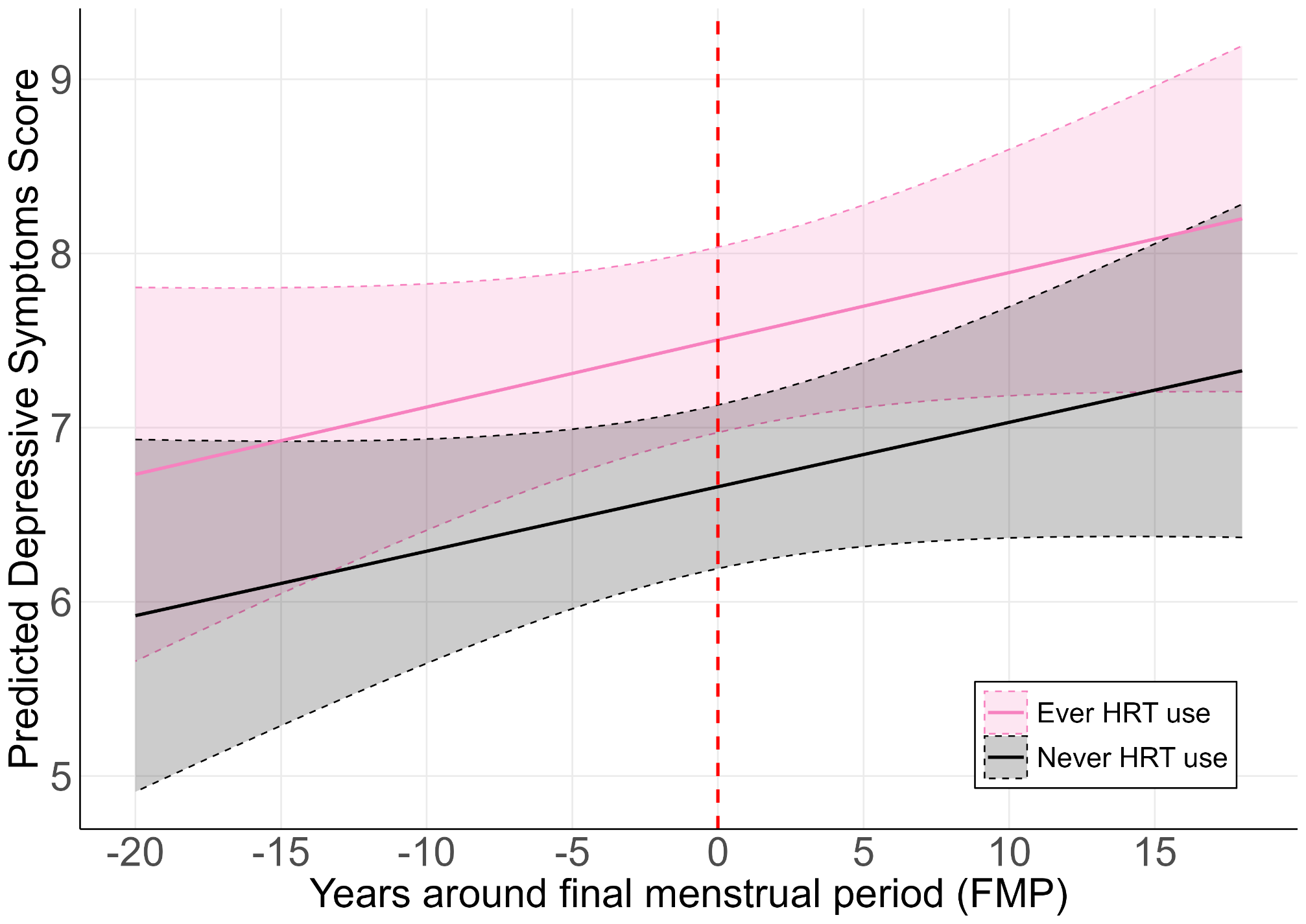
